# Simvastatin therapy in different subtypes of hypercholesterolemia – a physiologically based modelling approach

**DOI:** 10.1101/2023.02.01.23285358

**Authors:** Florian Bartsch, Jan Grzegorzewski, Helena Leal Pujol, Hans-Michael Tautenhahn, Matthias König

**Affiliations:** Institute for Theoretical Biology, Humboldt University, Berlin, Germany; Experimental Transplantation Surgery, Department of General, Visceral and Vascular Surgery, Jena University Hospital, Jena, Germany

**Keywords:** simvastatin, hypercholesterolemia, LDL-cholesterol, LDL-receptor, physiologically based pharmacokinetic model, pharmacokinetics, pharmacodynamics, PK/PD

## Abstract

Hypercholesterolemia is a multifaceted plasma lipid disorder with heterogeneous causes including lifestyle and genetic factors. A key feature of hypercholesterolemia is elevated plasma levels of low-density lipoprotein cholesterol (LDL-C). Several genetic variants have been reported to be associated with hypercholesterolemia, known as familial hypercholesterolemia (FH). Important variants affect the LDL receptor (LDLR), which mediates the uptake of LDL-C from the plasma, apoliporotein B (APOB), which is involved in the binding of LDL-C to the LDLR, and proprotein convertase subtilisin/kexin type 9 (PCSK9), which modulates the degradation of the LDLR. A typical treatment for hypercholesterolemia is statin medication, with simvastatin being one of the most commonly prescribed statins. In this work, the LDL-C lowering therapy with simvastatin in hypercholesterolemia was investigated using a computational modeling approach. A physiologically based pharmacokinetic model of simvastatin integrated with a pharmacodynamic model of plasma LDL-C (PBPK/PD) was developed based on extensive data curation. A key component of the model is LDL-C turnover by the liver, consisting of: hepatic cholesterol synthesis with the key enzymes HMG-CoA reductase and HMG-CoA synthase; cholesterol export from the liver as VLDL-C; de novo synthesis of LDLR; transport of LDLR to the membrane; binding of LDL-C by LDLR via APOB; endocytosis of the LDLR-LDL-C complex; recycling of LDLR from the complex. The model was applied to study the effects of simvastatin therapy in hypercholesterolemia due to different causes in the LDLR pathway corresponding to different subtypes of hypercholesterolemia. Model predictions of LDL-C lowering therapy were validated with independent clinical data sets. Key findings are: (i) hepatic LDLR turnover is highly heterogeneous among FH classes; (ii) despite this heterogeneity, simvastatin therapy results in a consistent reduction in plasma LDL-C regardless of class; and (iii) simvastatin therapy shows a dose-dependent reduction in LDL-C. Our model suggests that the underlying cause of hypercholesterolemia does not influence simvastatin therapy. Furthermore, our model supports the treatment strategy of stepwise dose adjustment to achieve target LDL-C levels. Both the model and the database are freely available for reuse.

## INTRODUCTION

Cholesterol is one of the most important and highly decorated molecules in biology (Brown et al., 1986). It is a critical structural component of cell membranes (Luo et al., 2019) and a precursor for a variety of important biomolecules such as bile acids, vitamin D or steroids (Cornforth and Popjaák, 1958). Cholesterol is a highly lipophilic compound and is transported in plasma by lipoproteins such as HDL (high density lipoprotein), LDL (low density lipoprotein), and VLDL (very low density lipoprotein).

The liver is the major site of cholesterol synthesis, clearance, and export into the plasma via VLDL cholesterol. 70% of the clearance of plasma LDL-C is mediated by LDL receptors on the membrane surface of hepatocytes (Gidding et al., 2015). Hepatic cholesterol levels can be increased by dietary cholesterol intake, uptake from plasma LDL-C, or hepatic *de novo* synthesis, and decreased by export to plasma, conversion to bile acids, or fecal excretion (Luo et al., 2019). The LDLR life cycle in the liver is tightly regulated by several key processes. These are (i) *de novo* synthesis of LDLR; (ii) transport of LDLR to the membrane; (iii) binding of extracellular LDL-C to membrane-bound LDLR via apolipoprotein B (APOB); (iv) endocytosis of the LDLR-LDL-C complex; and (v) recycling of LDLR from the complex. LDLR production rates in the liver are controlled by cholesterol levels in hepatocytes via negative feedback (Gidding et al., 2015).

A common abnormality in whole-body cholesterol homeostasis is elevated plasma levels of total cholesterol in combination with elevated LDL-C, known as hypercholesterolemia (Ibrahim et al., 2020). Hypercholesterolemia is associated with an increased risk for atherosclerosis, which can lead to cardio-, cerebro-, and peripheral morbidity and mortality (Christians et al., 1998). Cholesterol levels have become an important indicator of increased risk for cardiovascular disease (CVD) (Backer, 2003).

The main treatment for hypercholesterolemia is lifestyle changes such as lipid-lowering diets or lipid-lowering medications (Ibrahim et al., 2020). The most prominent medications are statins, which account for 95.8% of prescribed lipid-lowering medications in 2019 (Cheema et al., 2022). Statins can be used as lipid-lowering drugs because of their ability to inhibit the enzyme HMG-Coenzyme-A reductase (HMG-CoA reductase). This enzyme catalyzes the conversion of HMG-CoA to cholesterol, which is one of the major rate-limiting steps in cholesterol biosynthesis (Corsini et al., 1995). The resulting decrease in *de novo* cholesterol biosynthesis and hepatic cholesterol concentration leads to an upregulation of LDLR expression. As a consequence, the clearance of LDL-C from plasma by LDLR-mediated uptake into the liver increases, leading to a decrease in plasma LDL-C and total cholesterol concentrations (Magot et al., 1991).

Simvastatin is one of the most popular and widely used statins. In 2011-2012, 23.2% of adults aged 40 and over in the United States used simvastatin as a cholesterol-lowering medication (Gu, 2014). Simvastatin itself is a prodrug that must be activated in the liver by esterases to the main active metabolite, simvastatin acid. Both simvastatin and simvastatin acid are metabolized by cytochrome P450 3A (CYP3A4) in the small intestine and liver to other simvastatin metabolites that have less inhibitory activity than simvastatin acid. Simvastatin is a nonpolar and highly lipophilic compound capable of passive diffusion across biomembranes. After metabolism, the metabolites gain polarity and are subject to specific transporters (e.g., the hepatic influx of simvastatin acid is mediated by the OATP1B1 transporter) (Jiang et al., 2017). The rapid first-pass metabolism and hydrophilicity of simvastatin result in a bioavailability of only 5% (Mauro, 1993). These effects lead to accumulation of simvastatin acid and other active metabolites in the liver where they competitively inhibit HMG-CoA reductase (Germershausen et al., 1989). Simvastatin undergoes enterohepatic circulation and is excreted as a variety of different simvastatin metabolites predominantly in the feces.

Hypercholesterolemia is a multifaceted disease with heterogeneous causes. These include mainly genetic and lifestyle factors (Ibrahim et al., 2020). Genetic factors typically lead to familial hypercholesterolemia (FH), which describes elevated plasma LDL cholesterol levels due to genetic disorders affecting the function of LDL-R, apolipoprotein B (APOB), and proprotein convertase subtilisin/kexin type 9 (PCSK9) (Di Taranto et al., 2020). The different genetic variants can be classified as loss of function of LDLR or loss of binding capacity of LDLR to APOB on LDL-C particles (Di Taranto et al., 2020). FH can be classified into six classes (Gidding et al., 2015; Hobbs et al., 1992; Defesche et al., 2017):

- Class 1: LDLR or precursors are not synthesized.
- Class 2: LDLR is not properly transported from the endoplasmic reticulum to the Golgi apparatus for expression on the cell surface.
- Class 3: LDLR does not properly bind LDL because of a defect in either APOB-100 or in LDLR.
- Class 4: LDLR bound to LDL-C does not properly cluster in clathrin-coated pits for receptor-mediated endocytosis.
- Class 5: LDLR is not recycled back to the cell surface and is rapidly degraded.
- Class 6: LDLR is not initially transported to the basolateral membrane.

All classes result in reduced LDL-C uptake from plasma but have different underlying causes in the LDLR lifecycle (binding, uptake, recycling, synthesis, activity). These six classes are grouped into two main categories: (i) receptor-negative mutations, which result in no LDLR synthesis or the synthesis of a non-functional LDLR, and (ii) receptor-defective mutations, which result in the synthesis of a less effective LDLR (Defesche et al., 2017). In contrast to the six classes of FH, mutations in PCSK9 do not result in loss of function of proteins involved in LDLR expression, but in gain of function of proteins involved in LDLR degradation (Di Taranto et al., 2020).

An open question is how the different classes of FH affect lipid-lowering therapy with statins. A better understanding of simvastatin therapy and whether and how different treatment regimens work in the different classes may allow a more personalized approach to cholesterol-lowering therapy based on subgroup stratification. Because of the complex regulation of cholesterol synthesis and homeostasis and the pharmacokinetics and metabolism of simvastatin, this question has been difficult to answer. In addition, the correct dosage of simvastatin is a major challenge, as too low concentrations result in ineffectiveness (no or insufficient reduction in LDL-C) and too high concentrations result in possible side effects such as hypertension and muscle damage. It is unclear which dosing strategy should be used in which class. Computational modeling can be used as an important method to study such complex systems *in silico*.

Computational models of simvastatin (Moon and Smith, 2002; Ogungbenro et al., 2019; Tsamandouras et al., 2014, 2015; Kim et al., 2011; Methaneethorn et al., 2014; Lohitnavy et al., 2015; Kim et al., 2011; Wojtyniak et al., 2021), cholesterol (Paalvast et al., 2015; Wrona et al., 2015), and models of the effect of simvastatin on cholesterol levels (Kim et al., 2011) have been developed. Most of these studies used small patient cohorts for model development (mostly a single clinical trial) and lack validation with independent data sets. These models lack general applicability. Modeling approaches such as network analysis (Moon and Smith, 2002; Wojtyniak et al., 2021) or population pharmacokinetic models using mostly one-compartment models (Kim et al., 2011; Ogungbenro et al., 2019; Tsamandouras et al., 2014, 2015; Methaneethorn et al., 2014) for simvastatin work well to describe a given data set, but they often lack general interpretability because they do not explicitly represent physiology. They often include only simvastatin and sometimes simvastatin acid, ignoring other important active metabolites that contribute to the lipid-lowering effect of simvastatin. Such reduced models do not take into account how physiological changes, such as anthropometric factors, affect simvastatin therapy. One study used a large data set for model building and validation and examined the effect of simvastatin in combination with various drug-drug and drug-gene interactions (Wojtyniak et al., 2021). Importantly, different FH classes in relation to simvastatin therapy are not investigated in any of the existing models.

The aim of this work was to develop a physiologically based model of simvastatin and cholesterol to study LDL-C lowering therapy with simvastatin in different FH classes. Such a model can help to better understand the quantitative and qualitative effects of simvastatin treatment and its efficacy in patients with elevated plasma cholesterol. In addition, this model can be used to answer the open question of whether genetic screening could be beneficial for personalized simvastatin therapy, e.g. to adjust dosing protocols according to individual physiology and genetics.

## MATERIALS AND METHODS

### Data curation

A database of simvastatin pharmacokinetics and LDL-C pharmacodynamics in simvastatin therapy was established for model development and validation (see Tab. 1). The data set consists of concentration-time curves and pharmacokinetic parameters for simvastatin and its metabolites simvastatin acid, active and total HMG-CoA reductase inhibitors. The pharmacodynamic data set consists of time courses of plasma LDL-C concentration during simvastatin therapy.

Inclusion criteria for simvastatin studies were that the studies reported pharmacokinetics and/or concentration-time curves of simvastatin and its metabolites after single or multiple doses of simvastatin. Priority was given to studies reporting data under control conditions (healthy subjects without the intervention of other drugs). Inclusion criteria for cholesterol studies were that the studies reported plasma lipid concentrations or changes (at least for LDL-C) after single or multiple doses of simvastatin. For inclusion, studies had to report baseline concentrations before treatment. The data are accompanied by metadata about the subjects and groups studied (e.g., type of atherosclerosis) and the intervention used (e.g., dose and route of simvastatin administration). All data have been curated using an established curation pipeline (Grzegorzewski et al., 2022) and are available via the PK-DB pharmacokinetics database (https://pk-db.com) (Grzegorzewski et al., 2021).

### Computational model

A physiologically based PK/PD model was developed to predict simvastatin pharmacokinetics and cholesterol pharmacodynamics. The model is available in the Systems Biology Markup Language (SBML) (Hucka et al., 2019; Keating et al., 2020) under a CC-BY 4.0 license from https://github.com/matthiaskoenig/simvastatin-model. This paper used version 0.9.1 of the model (König and Bartsch, 2023). The model was developed using sbmlutils (Kö nig, 2022), simulated using sbmlsim (Kö nig, 2021) with libroadrunner (Somogyi et al., 2015; Welsh et al., 2023) as the high performance simulator, and visualized using cy3sbml (König et al., 2012).

### Model parameterization

For model calibration, literature values were used for physiological and kinetic parameters such as Michaelis-Menten constants, inhibition constants, reference concentration values, blood flows and tissue volumes (see Tab. S1). The remaining model parameters were fitted by minimizing the residuals between the concentration-time curves from the curated data and the model predictions. The subset of data used for parameter fitting is listed in Tab. 1. All single-dose simvastatin studies were used for parameter fitting, with the exception of Keskitalo2008 (Keskitalo et al., 2008), which was excluded from parameter fitting due to genetic variants. The optimization problem was formulated as a nonlinear, bounded-variable least-squares problem and solved using SciPy’s least-squares method to minimize residuals between model predictions and data points (Virtanen et al., 2020). A total of 17 model parameters related to simvastatin were fitted (see Tab. S2).

### Familial hypercholesterolemia subtypes

To study the effect of different subtypes of familial hypercholesterolemia on plasma LDL-C levels and hepatic cholesterol metabolism, model parameters have been added that allow the degree of functional change within different steps of the LDLR pathway to be scanned. These have a default value of 1 and can be scanned to simulate different classes of FH. These scaling parameters are listed in Tab. S3. FH classes 1, 3, 4, and 5 are implemented with a single scaling parameter. Both class 2 and class 6 affect the membrane transport of LDLR: class 2 affects transport from the ER to the Golgi apparatus for cell surface expression, and class 6 affects the initial transport of LDLR to the basolateral membrane. Membrane transport of LDLR was modeled by a single overall reaction that combined these steps. Consequently, class 2 and 6 could not be distinguished in the model and were described by a single parameter. To describe the effects of mutations in PCSK9, an additional scaling parameter affecting the degradation of LDLR was added. All parameters for the FH classes are set to 1.0 in the model reference state corresponding to 3 mM plasma LDL-C. Parameters have been varied in [0.01, 100] corresponding to different degrees of function.

### Baseline LDL-C values

The model was calibrated to a baseline reference plasma LDL-C of 3 mM. For calibration, LDL-C consumption was adjusted using the calibration curve in Fig. S1. To establish specific baseline plasma LDL-C concentrations in the different hypercholesterolemic classes, each FH parameter was adjusted using the calibration curves in Fig. S2. To determine these curves, each FH parameter was scanned in the range [10E-3, 10E3] and a time course simulation was performed over 52 weeks until steady-state LDL-C values were reached. The values were interpolated and the interpolation curve was used to determine the change in FH parameter for a given LDL-C value.

## RESULTS

In this work, we developed a physiologically based pharmacokinetic/pharmacodynamic (PBPK/PD) model of simvastatin (SV) and its pharmacodynamic effect on plasma LDL-C. The model was applied to study the LDL-C-lowering effect of simvastatin therapy in different subtypes of hypercholesterolemia.

### Database on the pharmacokinetics and pharmacodynamics of simvastatin

A large pharmacokinetic database of simvastatin and its metabolites (Tab. 1) combined with a pharmacodynamic database of the LDL-C lowering effect of simvastatin therapy (Tab. 2) was established for the development and evaluation of the model.

The pharmacokinetic database consists of 27 studies reporting simvastatin time courses or pharmacokinetic parameters. Of the studies, 23 report simvastatin (SV) time courses after single oral administration (Backman et al., 2000; Chung et al., 2006; Gehin et al., 2015; Jacobson, 2004; Jiang et al., 2017; Kantola et al., 1998; Keskitalo et al., 2008, 2009; Kim et al., 2019; Kyrklund et al., 2000; Lilja et al., 2000, 2004; Lohitnavy et al., 2004; Marino et al., 2000; Mousa et al., 2000; Neuvonen et al., 1998; Pasanen et al., 2006; Pentikainen et al., 1992; Tubic-Grozdanis et al., 2008; Ucar et al., 2004; Zhou et al., 2013). In addition, 7 studies report time courses after multiple oral administrations of simvastatin (Bergman et al., 2004; Hsyu et al., 2001; Jacobson, 2004; Nishio et al., 2005; Simard et al., 2001; Zhi et al., 2003; Ziviani et al., 2001). A single study reported both, time courses after single and multiple doses of simvastatin (Jacobson, 2004). 24 studies reported time courses for SV, 19 for simvastatin acid (SVA), 3 for SV + SVA, 5 for active HMG-CoA reductase inhibitors, and 5 for total HMG-CoA reductase inhibitors.

The pharmacodynamics data set consists of 18 studies that reported on plasma LDL-C or changes in plasma LDL-C with simvastatin therapy (see Tab. 2) (Crouse 3rd et al., 1999; Davidson et al., 1997; Geiss et al., 2002; Isaacsohn et al., 2003; Jones et al., 1998; Keech et al., 1994; Kosoglou et al., 2002; Loria et al., 1994; Li et al., 2003; Mö lgaard et al., 1988; Mol et al., 1986, 1988; Ntanios et al., 1999; Nishio et al., 2005; Owens et al., 1991; Pietro et al., 1989; Recto et al., 2000; Tuomilehto et al., 1994; Walker et al., 1990). Study duration was heterogeneous, ranging from 2 weeks to 3 years, with one study reporting plasma LDL-C after a single dose application (Loria et al., 1994). Most studies did not provide sufficient information on FH phenotypes in the study cohort. Some studies included subjects with polygenic, heterozygous, homozygous, mixed, or type 2 hypercholesterolemia. All study cohorts consisted of patients with elevated plasma LDL-C levels, except for Loria1993 (Loria et al., 1994).

To our knowledge, this is the first large freely available data set of pharmacokinetic and pharmacodynamic data for simvastatin with all data accessible from the pharmacokinetic database (PK-DB) (Grzegorzewski et al., 2021).

### PBPK/PD model of simvastatin

A physiologically based pharmacokinetic/pharmacodynamic (PBPK/PD) model of simvastatin (SV) and its pharmacodynamic effect on plasma LDL-C was developed to study SV therapy in different classes of hypercholesterolemia (Fig. 1). The whole-body model (Fig. 1A) consists of the liver, kidney, gastrointestinal tract, lungs, and the rest compartment. Organs of minor relevance are not explicitly modeled and are lumped into the rest compartment. Organs are coupled via the systemic circulation. SV can be administered orally (PO).

**Figure 1.**
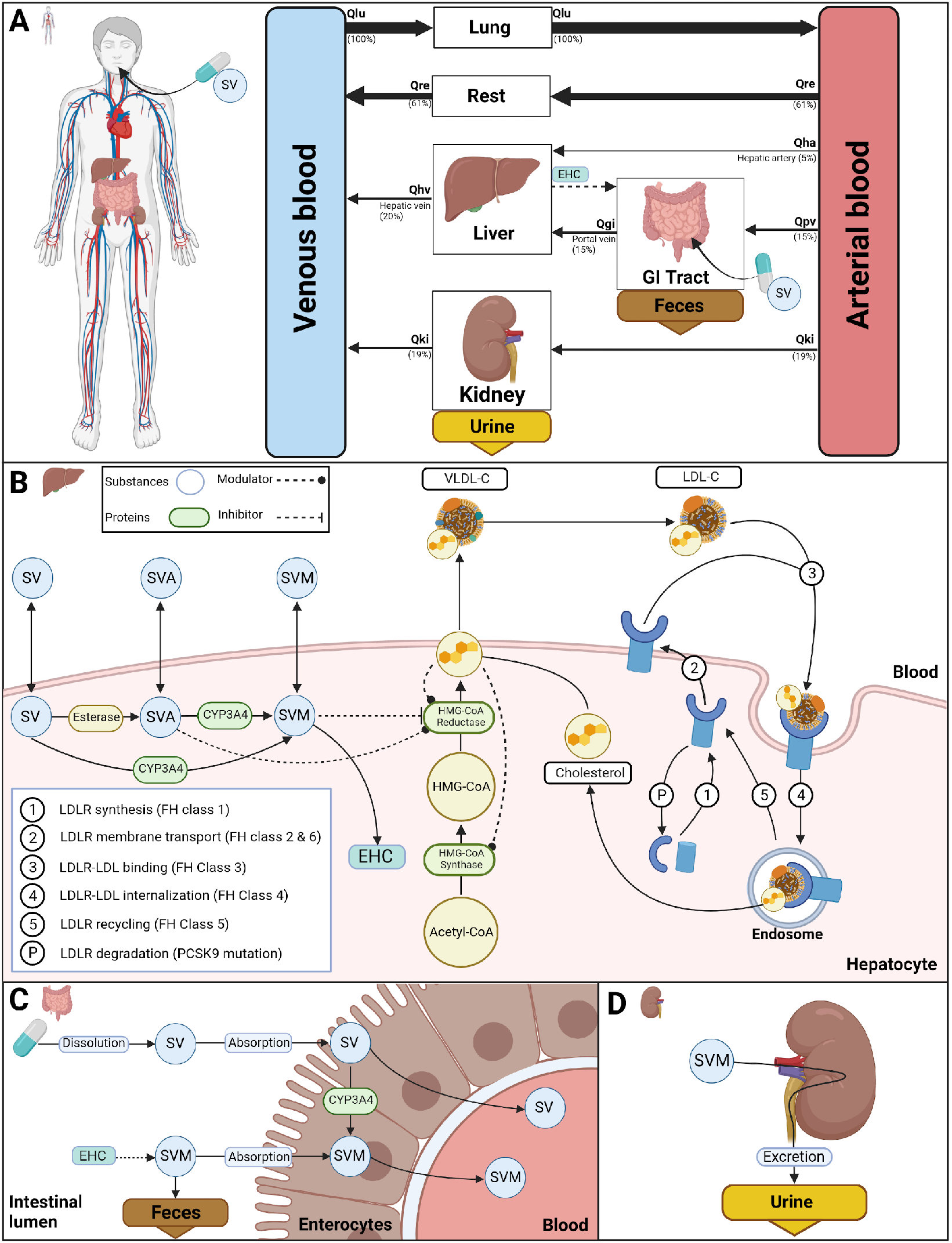
Physiologically based model of simvastatin and cholesterol. **A)** Whole-body model consisting of lung, liver, kidney, gastrointestinal tract and blood compartments. Simvastatin (SV), simvastatin acid (SVA), simvastatin metabolites (SVM), LDL cholesterol (LDL-C) and VLDL cholesterol (VLDL-C) are transported via the systemic circulation. **B)** Liver submodel including simvastatin metabolism, cholesterol synthesis and the LDL receptor (LDLR) pathway. SVA and SVM competitively inhibit HMG-CoA reductase. Cholesterol has a negative feedback on HMG-CoA reductase and HMG-CoA synthase. The LDLR pathway consists of: (1) synthesis of LDLR; (2) transport of LDLR to the membrane; (3) binding of LDL-C to LDLR; (4) internalization of the LDLR-LDL-C complex; (5) recycling of LDLR; and (P) degradation of LDLR. The liver exports cholesterol via VLDL-C particles and SVM into the bile, resulting in the enterohepatic circulation (EHC) of SVM. **C)** Submodel of the gastrointestinal tract including first-pass metabolism of SVM, enterohepatic circulation (EHC), and fecal excretion. SVM can reach the intestine via biliary transport from the liver. SV and SVM can be absorbed into the intestine via enterocytes. Within the enterocytes, SV is converted to SVM by CYP3A4. SV and SVM are transported into the blood. **D)** Kidney submodel consisting of urinary excretion of SVM via the kidneys. Created with BioRender.com.

In the liver model (Fig. 1B), SV is converted to SVM by CYP3A. Alternatively, esterases catalyze the reaction of SV to SVA with subsequent conversion to SVM by CYP3A4. This is the main activation process of simvastatin. SV, SVA, and SVM can be exchanged between the liver and the circulation. SVM accounts for all simvastatin metabolites after metabolism by either CYP3A4 or esterases. SVM can be transported via enterohepatic circulation (EHC) from the liver into the gastrointestinal tract.

The intestinal model (Fig. 1C) describes the dissolution and absorption of SV, absorption of SV, and first pass metabolism of SV to SVM via CYP3A4 in the enterocytes of the intestinal wall. Only a fraction of SVM that reaches the intestine via the enterohepatic circulation (EHC) is absorbed, with the remainder excreted in the feces, whereas SV can be completely absorbed.

The kidney model (Fig. 1D) describes the urinary excretion of SVM. There is no renal clearance of SV and SVA.

To study the effects of simvastatin therapy, the simvastatin pharmacokinetic model was extended to include the major processes affecting plasma LDL-C levels. The liver model (Fig. 1B) includes the major processes relevant to hepatic cholesterol homeostasis, including dietary cholesterol uptake, fecal cholesterol loss, a shortened cholesterol biosynthetic pathway, and uptake of cholesterol from plasma as LDL-C. Cholesterol can be exported from the liver as VLDL-C. Hepatic cholesterol synthesis is modeled via the precursor reaction from acetyl-CoA to HMG-CoA mediated by HMG-CoA synthase and the reaction from HMG-CoA to cholesterol mediated by HMG-CoA reductase. SVA and SVM are competitive inhibitors of HMG-CoA reductase. An important regulatory mechanism in the model is the adjustment of protein levels of HMG-CoA synthase, HMG-CoA reductase and LDLR by hepatic cholesterol levels. As cholesterol levels decrease, protein synthesis rates increase and protein degradation rates decrease for these proteins, resulting in an increase in these key enzymes of cholesterol synthesis.

A key component of the model is the hepatic LDL receptor (LDLR) pathway consisting of (1) LDLR synthesis; (2) transport of LDLR to the membrane; (3) binding of LDL-C to LDLR; (4) internalization of the LDLR-LDL-C complex; (5) recycling of LDLR; and (P) degradation of LDLR. The effect of altering these steps was systematically evaluated to implement different subtypes of familial hypercholesterolemia.

To our knowledge, this is the first freely available, reproducible, and reusable PBPK/PD model of simvastatin pharmacokinetics and pharmacodynamics with the model available in SBML from https://github.com/matthiaskoenig/simvastatin-model.

### Simvastatin time courses

The performance of the simvastatin PBPK model was evaluated by comparing model predictions of the time courses of simvastatin and its metabolites with data from the curated studies. Time course data after a single dose of simvastatin were used as the training data set, and data after multiple doses of simvastatin were used as an independent validation data set (see Tab. 1). Model predictions for simvastatin, simvastatin acid, total simvastatin inhibitors, active simvastatin inhibitors and simvastatin plus simvastatin acid (see Fig. 2) were in good agreement with the training data from 21 studies (Backman et al., 2000; Chung et al., 2006; Gehin et al., 2015; Jacobson, 2004; Jiang et al., 2017; Kantola et al., 1998; Keskitalo et al., 2008, 2009; Kim et al., 2019; Kyrklund et al., 2000; Lilja et al., 2000, 2004; Lohitnavy et al., 2004; Marino et al., 2000; Mousa et al., 2000; Neuvonen et al., 1998; Pasanen et al., 2006; Pentikainen et al., 1992; Tubic-Grozdanis et al., 2008; Ucar et al., 2004; Zhou et al., 2013). Similar good performance of the model was observed on the validation data set consisting of 7 studies with good agreement between model predictions and data (see Fig. 2) (Bergman et al., 2004; Hsyu et al., 2001; Jacobson, 2004; Nishio et al., 2005; Simard et al., 2001; Zhi et al., 2003; Ziviani et al., 2001). The resulting simvastatin PBPK model accurately predicts plasma concentrations of simvastatin and its metabolites under single and multiple applications of simvastatin.

**Figure 2.**
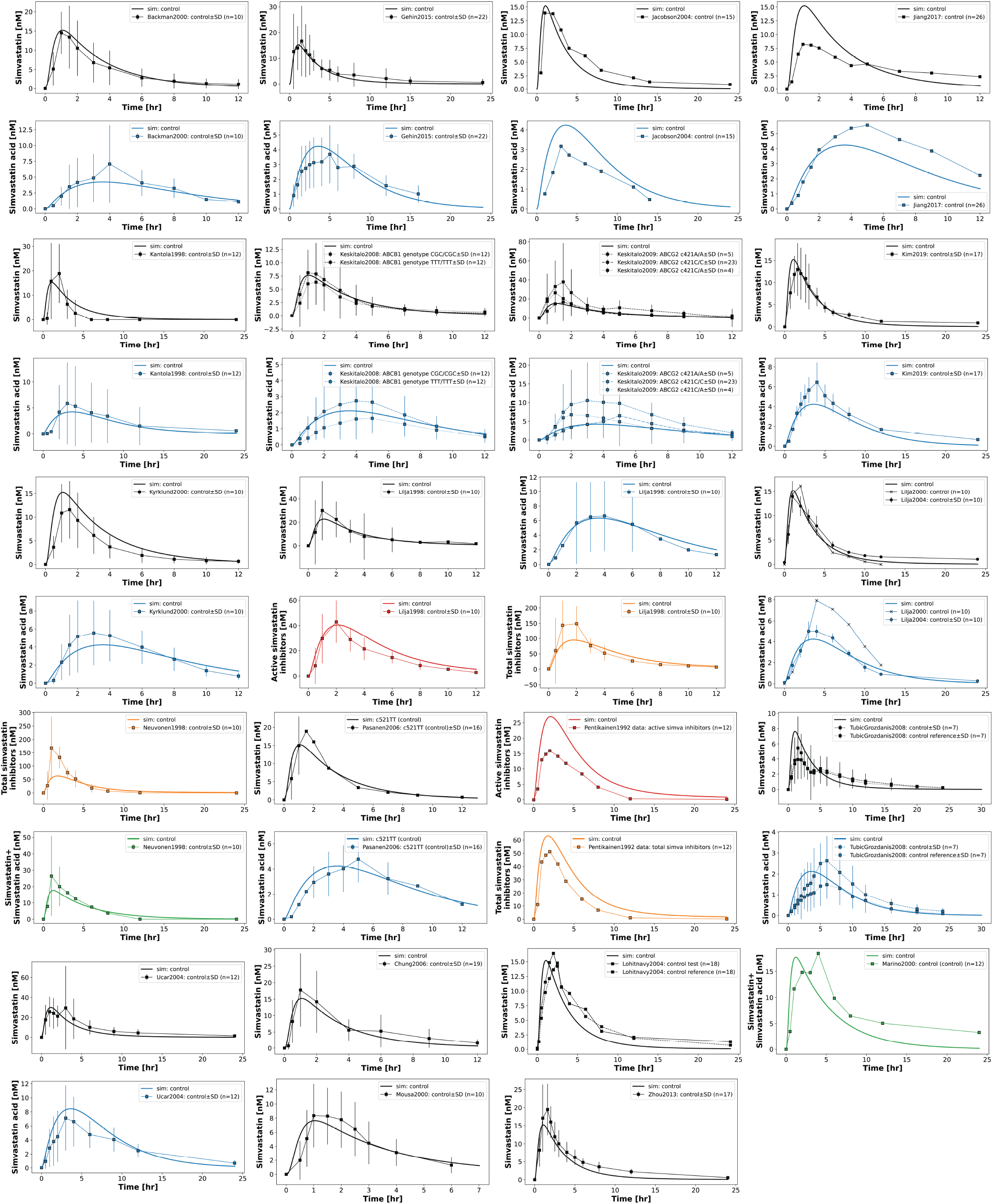
Time courses after single application of simvastatin. Simvastatin model performance on the training data set consisting of a single oral dose of simvastatin. Model predictions for plasma concentrations of simvastatin (black), simvastatin acid (blue), total simvastatin inhibitors (orange), active simvastatin inhibitors (red), and simvastatin plus simvastatin acid (green). Means are shown or mean *±*SD if SD was reported in the study. For the simulation, the oral dose was set according to the dosing protocol in Tab. 1. Data from (Backman et al., 2000; Chung et al., 2006; Gehin et al., 2015; Jacobson, 2004; Jiang et al., 2017; Kantola et al., 1998; Keskitalo et al., 2008, 2009; Kim et al., 2019; Kyrklund et al., 2000; Lilja et al., 2000, 2004; Lohitnavy et al., 2004; Marino et al., 2000; Mousa et al., 2000; Neuvonen et al., 1998; Pasan**1**en**9** et al., 2006; Pentikainen et al., 1992; Tubic-Grozdanis et al., 2008; Ucar et al., 2004; Zhou et al., 2013). Keskitalo2008 was not included in training of the model.

### Simvastatin therapy in hypercholesterolemia subtypes

The PBPK/PD model was calibrated to a baseline LDL-C concentration of 3 mM. To study the effect of modifying key steps of the LDLR pathway, the FH parameters for the respective familial hypercholesterolemia subtypes were varied by a factor of 10 at time 0 in the direction of increasing LDL-C levels. Time course simulations were performed for 52 weeks of no therapy followed by either simvastatin therapy with 20 mg simvastatin daily or no therapy for 52 weeks (Fig. 4). A zoom of the last week of simvastatin treatment is shown in Fig. S3. As a control in the no mutation simulation, no changes in the LDLR pathway were made under the same simvastatin dosing regimen.

Alterations in (1) LDLR synthesis (FH class 1), (2) transport of LDLR to the membrane (FH class 2 & 6), (3) binding of LDL-C to LDLR (FH class 3), (4) internalization of the LDLR-LDL-C complex (FH class 4), (5) recycling of LDLR (FH class 5), and (P) degradation of LDLR (PSCK9 mutation) all lead to hypercholesterolemia, but to different degrees (Fig. 4C). It takes 15-25 weeks for each FH class to reach a new LDL-C steady state after the change in the LDL-R pathway.

Simvastatin therapy reduces LDL-C in all subtypes by 2-4 mM. It takes approximately 20 weeks for each class to reach new LDL-C steady states after initiation of simvastatin therapy. Large daily fluctuations in metabolites and rates are observed during simvastatin therapy (Fig. S3). Both SV and SVA vary throughout the day with peaks around 2-3 hours after dosing (see also Fig. 2 and Fig. 3), with concentrations declining to almost zero after 24 hours due to the relatively fast half-life of simvastatin. As a consequence of the diurnal variation of HMG-CoA reductase inhibitors, the rate of hepatic cholesterol synthesis, hepatic cholesterol levels and the rate of VLDL-C export show large variations (Fig. 4D and H).

**Figure 3.**
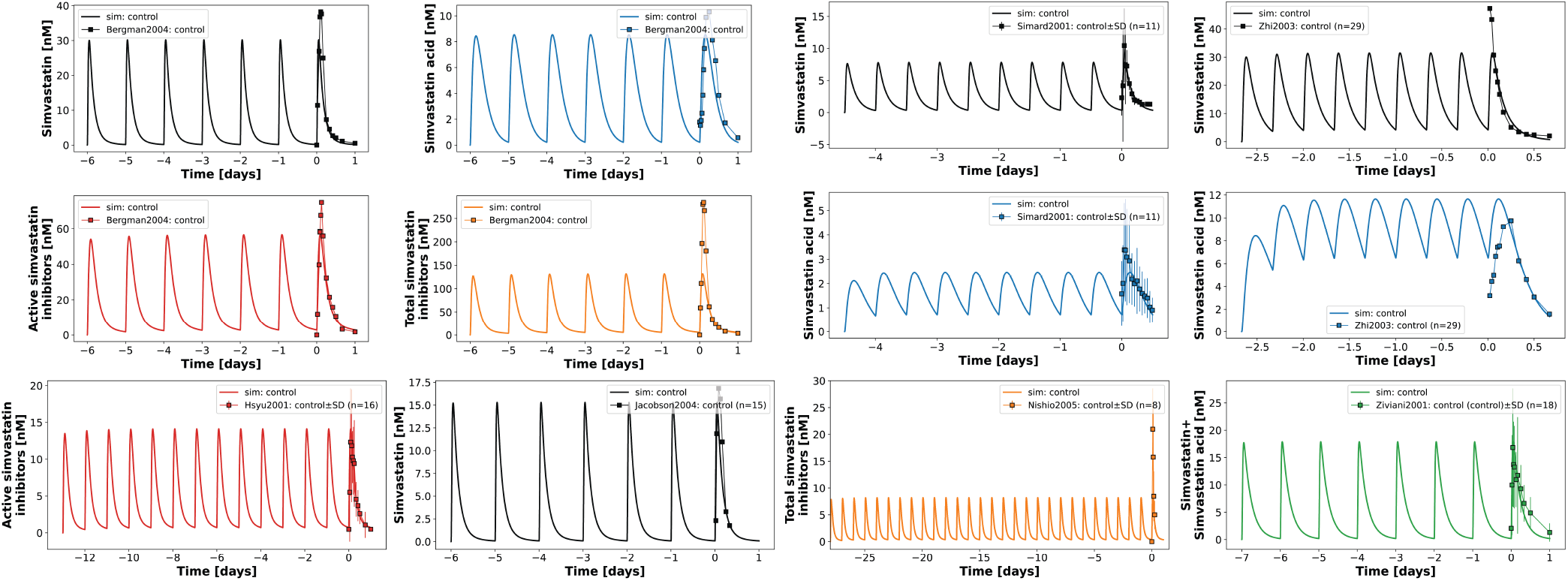
Time courses after multiple applications of simvastatin. Simvastatin model performance on validation data consisting of multiple oral dosing of simvastatin. Model predictions for simvastatin (black), simvastatin acid (blue), total simvastatin inhibitors (orange), active simvastatin inhibitors (red), and simvastatin plus simvastatin acid (green) after multiple oral doses of simvastatin. Means are shown or mean *±*SD if SD was reported in the study. For the simulation, oral doses were set according to the dosing protocol in Tab. 1.. Data from (Bergman et al., 2004; Hsyu et al., 2001; Jacobson, 2004; Nishio et al., 2005; Simard et al., 2001; Zhi et al., 2003; Ziviani et al., 2001). Data from Prueksaritanont et al. (2001) was excluded.

The different subtypes of hypercholesterolemia show a similar effect on LDL-C levels, consisting of an increase in LDL-C followed by a reduction with simvastatin therapy. Despite these similarities, large differences can be observed in the LDL-R pathway (Fig. 4E-I).

**Figure 4.**
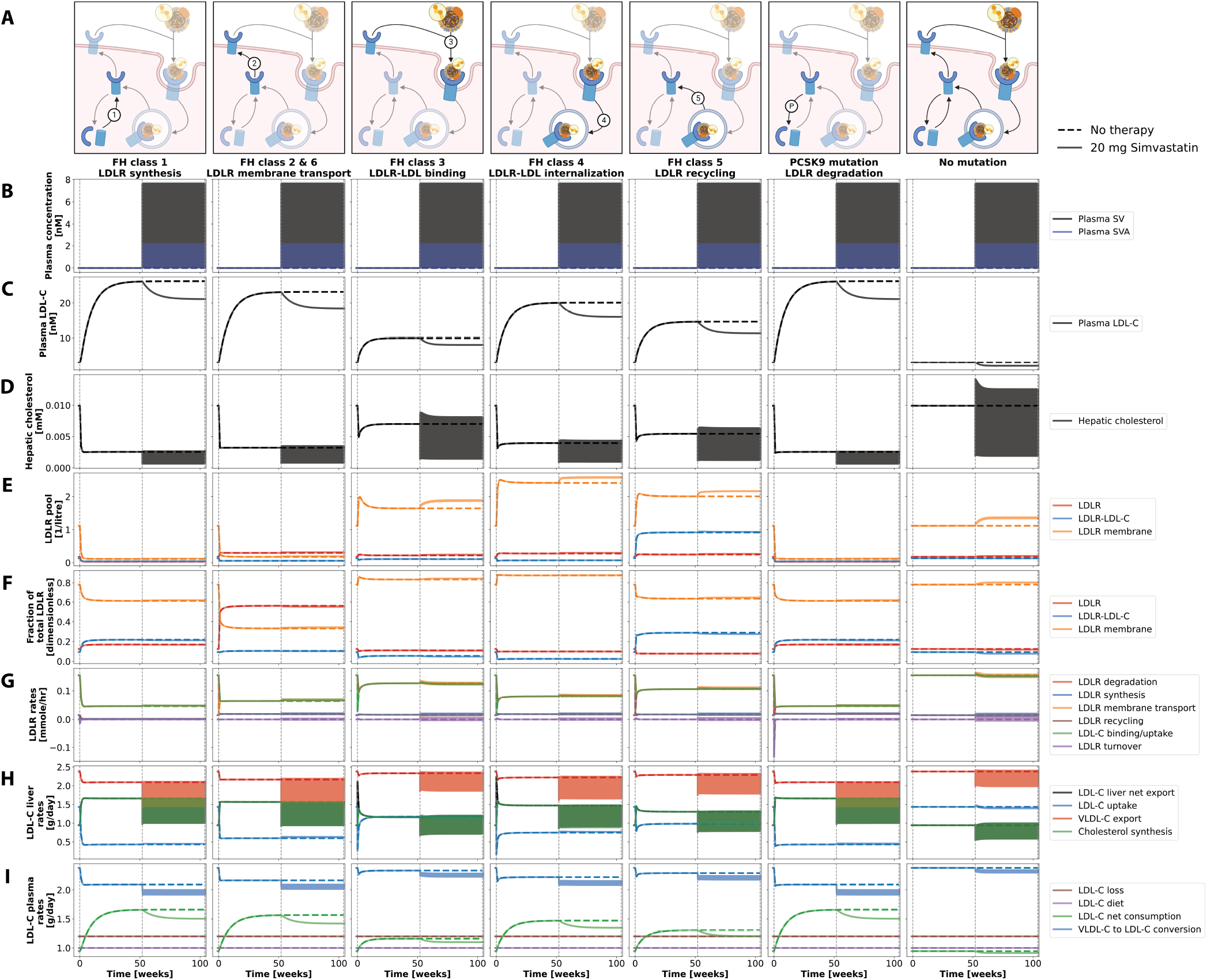
Time course simulation of hypercholesterolemia classes and simvastatin therapy. The columns correspond to the different classes of hypercholesterolemia. The simulation starts with a baseline reference value of 3 mM LDL-C, corresponding to no changes in the LDLR pathway. At time 0 weeks, the FH parameter is set to either 0.1 or 10, depending on the class, and simulated for 52 weeks, resulting in hypercholesterolemia. After 52 weeks, either 20 mg simvastatin daily for 52 weeks (solid lines) or no therapy (dashed lines) was applied. As a control, the no mutation simulation does not change any FH parameter. For a zoom on the last week, see Fig. S3. **A)** Graphic overview of the hypercholesterolemia subtypes: no mutation in the LDLR pathway; (1) LDLR synthesis; (2) transport of LDLR to the membrane; (3) binding of LDL-C to LDLR; (4) internalization of the LDLR-LDL-C complex; (5) recycling of LDLR; (P) degradation of LDLR; **B)** Plasma concentration of SV and SVA. **C)** Plasma LDL-C. **D)** Hepatic cholesterol. **E)** Overview of the LDLR pool consisting of plasma LDLR, LDLR-LDL-C complex or membrane LDLR. **F)** Fractional LDLR pool. **G)** Rate of processes involved in LDLR turnover: LDLR degradation, LDLR synthesis, LDLR membrane transport, LDLR recycling, LDL-C binding/uptake, LDL-R turnover. **H)** LDL-C rates in the liver: LDL-C net export from liver, LDL-C uptake, LDL-C absorption, LDL-C export, cholesterol synthesis. **I)** Plasma LDL-C Rates: LDL-C fecal loss, LDL-C from diet, LDL-C net consumption, VLDL-C to LDL-C conversion.

To study the effect of the degree of change in the hypercholesterolemia subtypes, the FH parameters were varied in [0.01, 100] (Fig. 5). Simulations were analogous to Fig. 4, i.e. 52 weeks of no therapy followed by either simvastatin therapy with 20 mg simvastatin daily or no therapy for 52 weeks. Values represent the mean *±* SD concentration of the last day.

**Figure 5.**
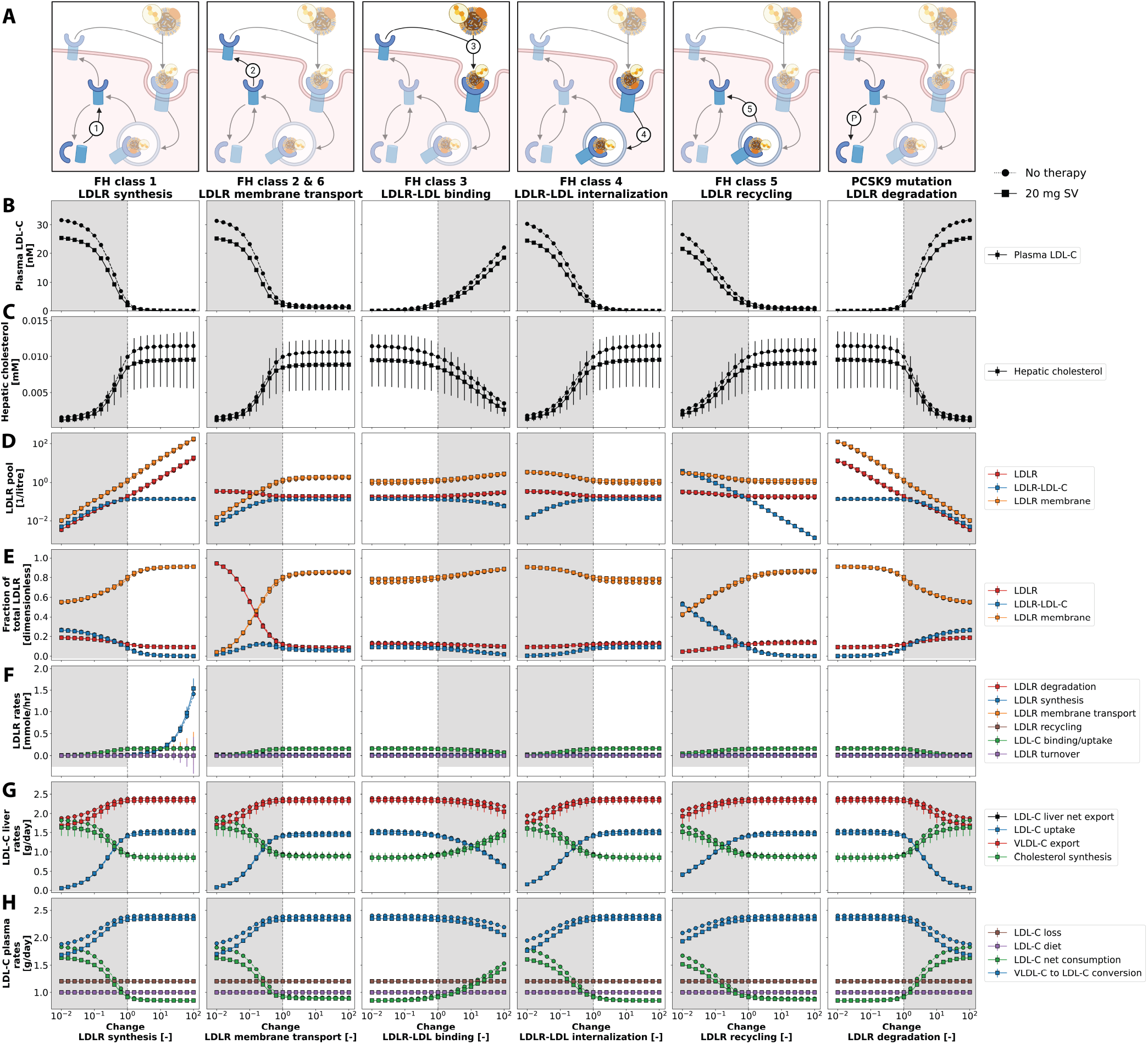
Steady-state simulation of hypercholesterolemia classes and simvastatin therapy. Simulations were performed as shown in Fig. 4 with FH parameters varied in [0.01, 100]. Shown are steady-state values after 52 weeks of either 20 mg simvastatin therapy (circle, solid lines) or no therapy (squares, dashed lines). Values correspond to the mean *±*SD daily concentration of the last day. Gray areas indicate the range of FH parameter changes that lead to hypercholesterolemia (LDL-C ¿ 3.0 mM). **A)** Graphic overview of the hypercholesterolemia subtypes: no mutation in the LDLR pathway; (1) LDLR synthesis; (2) transport of LDLR to the membrane; (3) binding of LDL-C to LDLR; (4) internalization of the LDLR-LDL-C complex; (5) recycling of LDLR; (P) degradation of LDLR; **B)** Plasma concentration of SV and SVA. **C)** Plasma LDL-C. **D)** Hepatic cholesterol. **E)** Overview of the LDLR pool consisting of plasma LDLR, LDLR-LDL-C complex or membrane LDLR. **F)** Fractional LDLR pool. **G)** Rate of processes involved in LDLR turnover: LDLR degradation, LDLR synthesis, LDLR membrane transport, LDLR recycling, LDL-C binding/uptake, LDL-R turnover. **H)** LDL-C rates in the liver: LDL-C net export from liver, LDL-C uptake, LDL-C absorption, LDL-C export, cholesterol synthesis. **I)** Plasma LDL-C Rates: LDL-C fecal loss, LDL-C from diet, LDL-C net consumption, VLDL-C to LDL-C conversion.

Plasma LDL-C levels show a sigmoidal dependence on the FH parameter, i.e. changes in key steps of the LDLR pathway are monotonically related to changes in plasma LDL-C levels. Plasma LDL-C does not vary much during the day. Interestingly, hepatic cholesterol behaves inversely to plasma LDL-C. High hepatic cholesterol concentrations result in low plasma LDL-C concentrations with large variations in hepatic cholesterol, and low hepatic cholesterol concentrations result in high plasma LDL-C concentrations with small variations over a day.

The different subtypes of hypercholesterolemia show a similar effect on LDL-C levels, consisting of an increase in LDL-C followed by a reduction with simvastatin therapy. Despite these similarities, large differences can be observed in the LDL-R pathway (Fig. 5E-I).

### Prediction of LDL-C reduction with simvastatin therapy

We then used the developed model to predict the reduction in plasma LDL-C with simvastatin therapy (Fig. 6) for data from (Crouse 3rd et al., 1999; Davidson et al., 1997; Geiss et al., 2002; Isaacsohn et al., 2003; Jones et al., 1998; Keech et al., 1994; Kosoglou et al., 2002; Loria et al., 1994; Li et al., 2003; Mölgaard et al., 1988; Mol et al., 1986, 1988; Ntanios et al., 1999; Nishio et al., 2005; Owens et al., 1991; Pietro et al., 1989; Recto et al., 2000; Saito et al., 1991; Tuomilehto et al., 1994; Walker et al., 1990). Simvastatin dose, dosing interval, and duration of therapy were used to simulate each study (see Tab. 2). Baseline LDL-C levels were adjusted individually for each simulation to match the data. For each study, all hypercholesterolemia subtypes were simulated and the mean and range of predictions were compared with the data.

**Figure 6.**
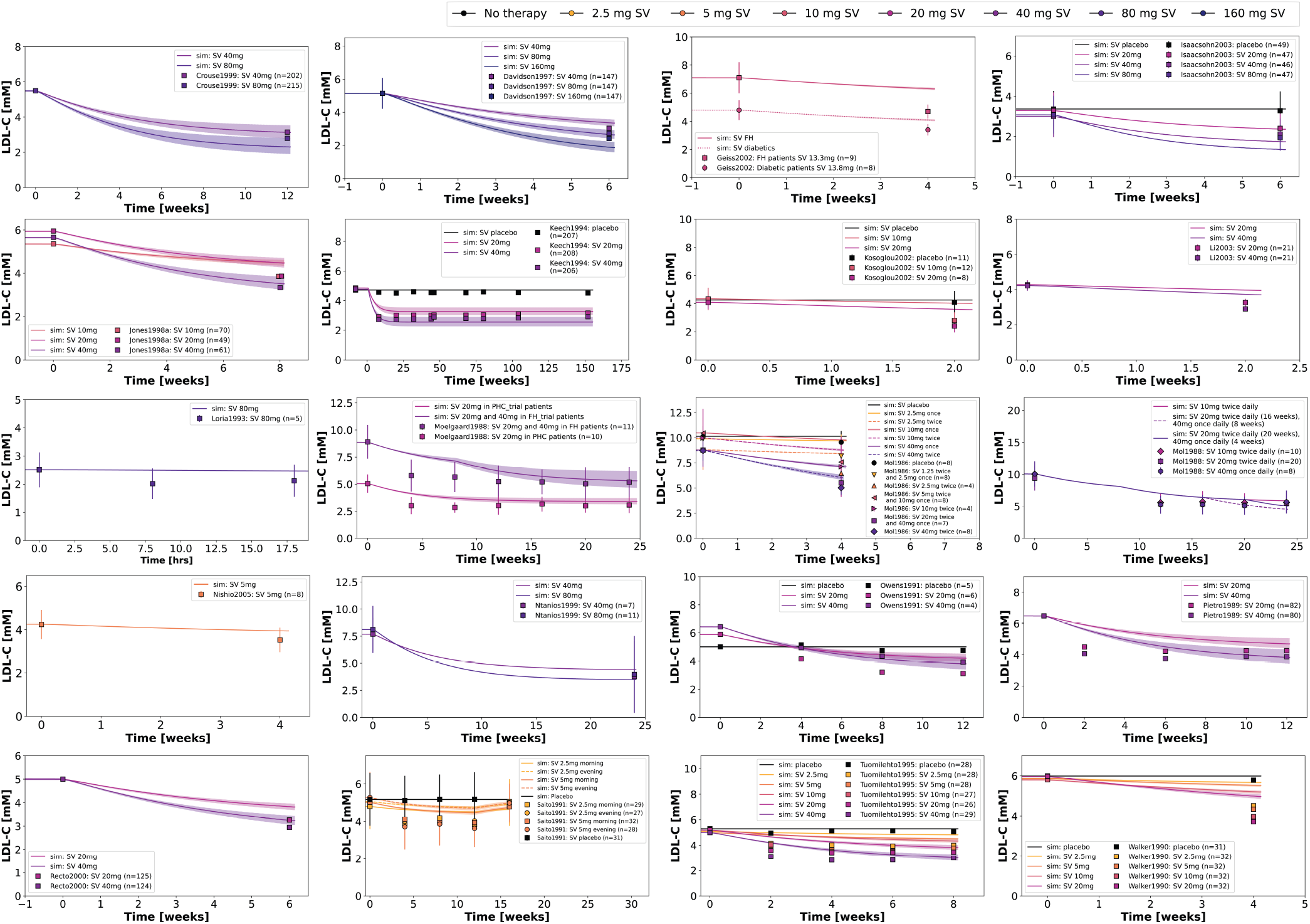
Prediction of LDL-C time course with simvastatin therapy. Time courses of plasma concentrations of LDL-C after multiple doses of oral simvastatin. For prediction, multiple model simulations were performed according to the different hypercholesterolemia classes using the dosing regimen of each study (see Tab. 2). For a single simulation, the respective FH parameter was adjusted to achieve the reported baseline LDL-C concentration. Simulation curves are the mean of the six FH classes with shaded areas corresponding to the range. The color corresponds to the respective dose of simvastatin. Data are mean or mean*±* SD when SD was reported. Data from (Crouse 3rd et al., 1999; Davidson et al., 1997; Geiss et al., 2002; Isaacsohn et al., 2003; Jones et al., 1998; Keech et al., 1994; Kosoglou et al., 2002; Loria et al., 1994; Li et al., 2003; Mölgaard et al., 1988; Mol et al., 1986, 1988; Ntanios et al., 1999; Nishio et al., 2005; Owens et al., 1991; Pietro et al., 1989; Recto et al., 2000; Saito et al., 1991; Tuomilehto et al., 1994; Walker et al., 1990).

Simvastatin therapy leads to a reduction in plasma LDL-C levels within a few weeks of treatment initiation. The predicted LDL-C time courses were generally in good agreement with the data over a wide range of simvastatin doses and dosing protocols. The model predictions are analyzed more systematically in Fig. 7 and Fig. S4. The predicted data are in good agreement with the observed data, especially for absolute reductions larger than -2 mM (Fig. 7A) and for relative reductions larger than -30% (Fig. 7D).

**Figure 7.**
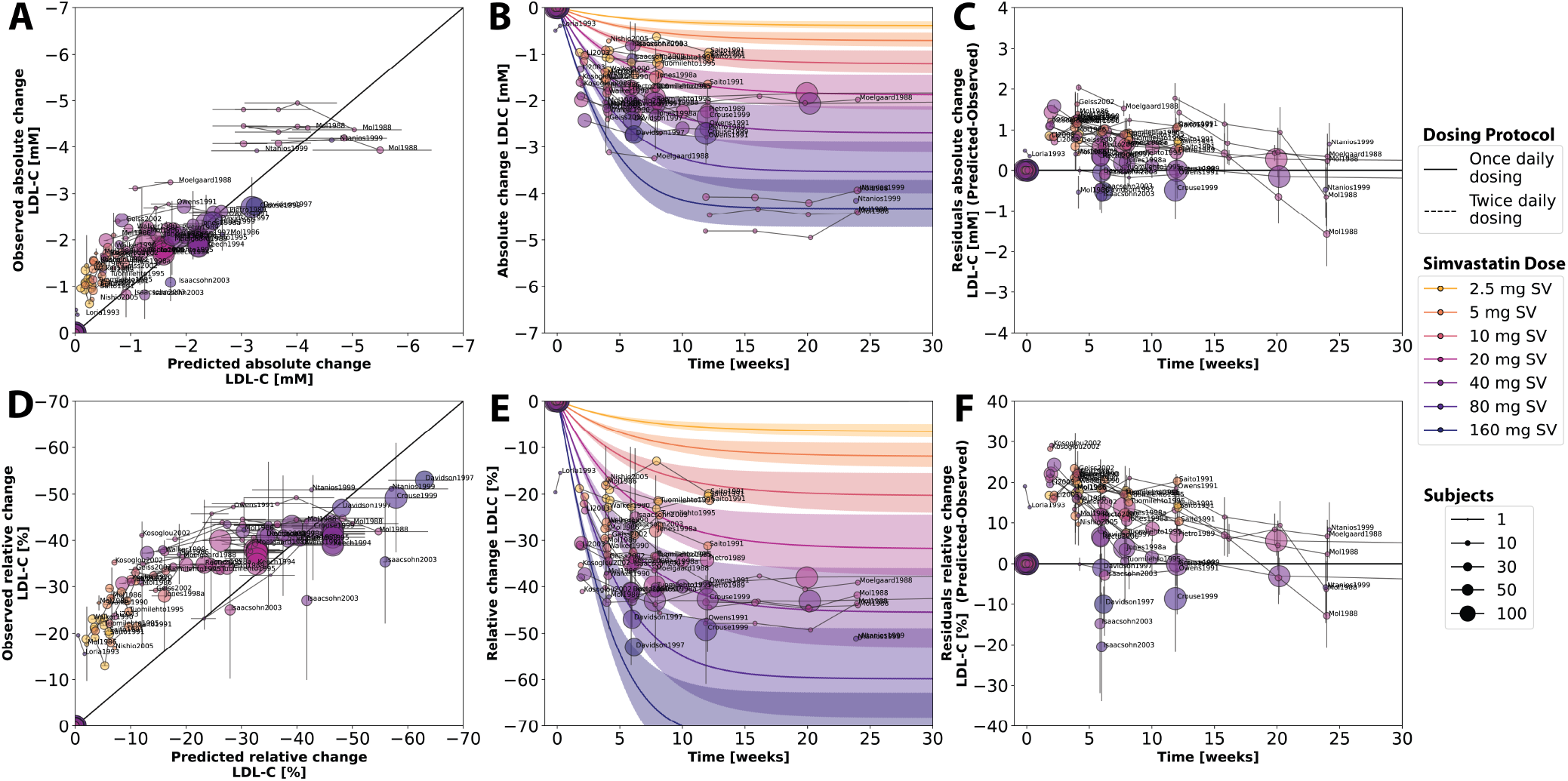
Prediction of LDL-C change with simvastatin therapy. Comparison of predicted and observed LDL-C changes with simvastatin therapy. Simulations and data from Fig. 6. **A)** Observed vs. predicted absolute change in LDL-C. Observed changes are mean SD, predicted changes are mean across FH classes with errors corresponding to the range. For changes for individual FH classes, see Fig. S4. **B)** Observed absolute changes in LDL-C over time with reference simulations. Observed changes are mean *±*SD. Simulations were initialized with a baseline plasma LDL-C value of 5.9 mM, which is the mean baseline value across all datasets. Simvastatin doses were applied every 24 h for 52 weeks. Lines are mean values across FH classes and shaded areas are minimum and maximum values across FH classes. **C)** Residuals of absolute observed changes and absolute predicted changes in plasma LDL-C with simvastatin therapy versus time. SD values were calculated using error propagation. **D)** Same as A but for relative changes in LDL-C. **E)** Same as B but for relative changes in LDL-C. **F)** Same as C) but for relative changes in LDL-C. Some studies did not report relative changes, only concentrations. Absolute and relative changes were calculated using the reported baseline LDL-C values. When studies reported only relative changes, absolute changes were calculated from relative changes with baseline values. SD values from relative changes were plotted and converted to SD for absolute changes using the coefficient of variation. For Keech1994, the reported baseline value at -8 weeks was used for time 0 weeks. Data from (Crouse 3rd et al., 1999; Davidson et al., 1997; Geiss et al., 2002; Isaacsohn et al., 2003; Jones et al., 1998; Keech et al., 1994; Kosoglou et al., 2002; Li et al., 2003; Loria et al., 1994; Mö lgaard et al., 1988; Mol et al., 1986, 1988; Nishio et al., 2005; Ntanios et al., 1999; Owens et al., 1991; Pietro et al., 1989; Recto et al., 2000; Saito et al., 1991; Tuomilehto et al., 1994; Walker et al., 1990).

To examine the time and dose dependence of LDL-C reduction, the data were compared to a reference simulation of the model with a baseline LDL-C of 5.9 mM. The simulations were performed for all hypercholesterolemic subtypes and the mean and range are shown for absolute LDL-C reductions in Fig. 7B and for relative LDL-C reductions from baseline in Fig. 7E. A clear dose-response relationship can be observed, with increasing simvastatin dose leading to increasing LDL-C reduction. After 15-20 weeks of therapy, the model reaches maximum LDL-C reductions. Finally, the corresponding time-dependent residuals were calculated for absolute LDL-C reductions in Fig. 7C and relative LDL-C reductions from baseline in Fig. 7F.

### Effect of simvastatin dose and hypercholesterolemia subtype on LDL-C reduction

As shown in Fig. 7, there is a clear dose-dependent effect in LDL-C reduction with simvastatin therapy. To systematically study this effect in different subtypes of hypercholesterolemia, simvastatin was administered at daily doses of 2.5 mg, 5 mg, 10 mg, 20 mg, 40 mg, 80 mg, and 160 mg. The resulting plasma LDL-C and absolute and relative reductions by subtype and magnitude of change are shown in Fig. 8. Simvastatin is able to reduce plasma LDL-C in each subtype (Fig. 8B). The absolute reduction (Fig. 8C) depends on the plasma LDL-C level, with higher pretreatment LDL-C levels resulting in greater absolute reductions. With increasing simvastatin dose, the absolute reductions and relative inductions increase. An important finding is that similar relative reductions are achieved at a given dose regardless of the underlying subtype of familial hypercholesterolemia and the extent of alteration in the LDLR pathways (Fig. 8D).

**Figure 8.**
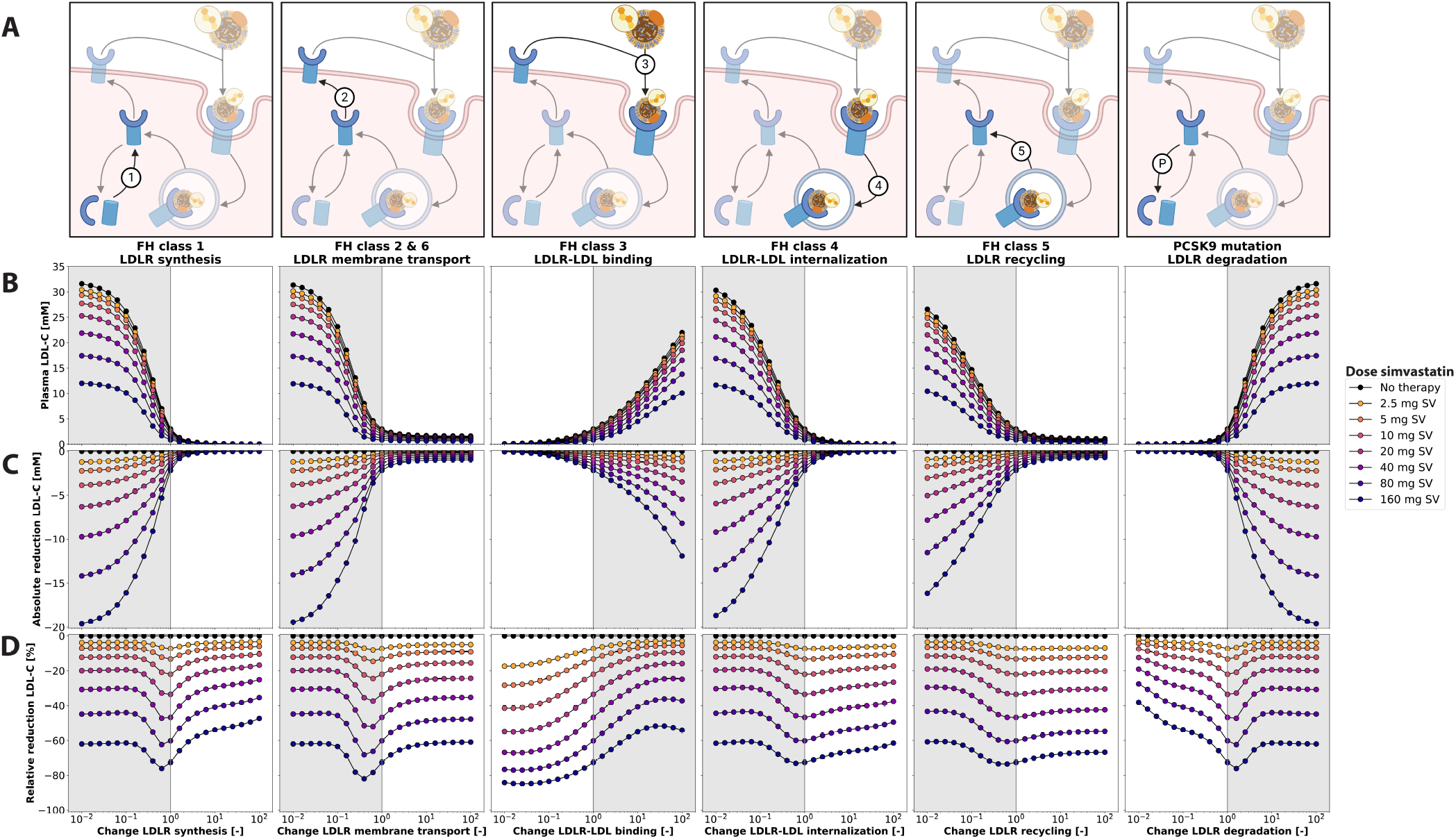
Relative and absolute LDL-C reductions depending on hypercholesterolemia class with simvastatin therapy. **A)** Graphic overview of the classes of hypercholesterolemia: (1) LDLR synthesis; (2) transport of LDLR to the membrane; (3) binding of LDL-C to LDLR; (4) internalization of the LDLR-LDL-C complex; (5) recycling of LDLR; (P) degradation of LDLR; **B)** Plasma LDL-C concentrations after 52 weeks of simvastatin therapy **C)** Absolute LDL-C reduction with simvastatin therapy versus no therapy. **D)** Relative LDL-C reduction versus no treatment. Gray areas indicate parameter ranges that lead to hypercholesterolemia (LDL-C *>* 3 mM). Simulation of 52 weeks of daily simvastatin therapy at various doses. Values are averaged from the last day of treatment. FH parameters were modified in [10E-3, 10E3]. The model was simulated for 59 weeks to reach steady state before treatment, followed by 52 weeks of therapy.

## DISCUSSION

In this work, a physiologically based pharmacokinetic/pharmacodynamic (PBPK/PD) model of simvastatin was developed and applied to study the LDL-C-lowering effect of simvastatin therapy in different subtypes of hypercholesterolemia.

The main findings of this paper are: (i) Hepatic LDLR turnover is highly heterogeneous among FH classes; despite the very similar effect of the different alterations on plasma LDL-C, i.e. increased LDL-C levels that could be reduced with simvastatin therapy, large differences in LDLR pathways and hepatic metabolites were observed among the different classes.

(ii) Despite this heterogeneity, simvastatin therapy results in a consistent lowering of plasma LDL-C regardless of class; the relative reduction for a given simvastatin dose is very similar regardless of the underlying cause. Our model suggests that inhibition of HMG-CoA reductase and cholesterol synthesis is an effective way to reduce elevated plasma LDL-C for all subtypes studied here. Our model suggests that the underlying cause of hypercholesterolemia in the FH classes does not affect simvastatin therapy. To answer the open question of whether genetic screening could be useful for personalized simvastatin therapy, e.g. to adjust dosing protocols according to individual physiology and genetics, our model says no.

Testing could provide valuable information for understanding the underlying individual pathophysiology of hypercholesterolemia, but will not add value to simvastatin therapy.

(iii) Simvastatin therapy shows a dose-dependent reduction in LDL-C. Our model supports the treatment strategy of stepwise dose adjustment to achieve target LDL-C levels. Both the model and the database are freely available for reuse.

The model spans time scales from very fast hepatic response kinetics to daily simvastatin therapy of up to years. The model incorporates slow adaptation processes that require 20-30 weeks after mutations in the LDLR pathway to reach steady state LDL-C levels. These slow timescales are also evident during simvastatin therapy, which requires a few weeks of daily dosing to achieve maximal reductions in plasma LDL-C. Daily peaks of HMG-CoA reductase inhibition are observed due to the rapid half-life of simvastatin, which reduces plasma concentrations to nearly zero after 24 hours. The short-term simvastatin pharmacokinetic model, coupled with a long-term cholesterol pharmacodynamic model, allows the effects of simvastatin to be studied in detail. Within the cholesterol model, we have been able to relate short-term inhibition of synthesis to long-term adjustments in protein levels. This allows us to predict and compare simvastatin-lowering therapy in patients with different FH types on a timescale of seconds to years. The model can discriminate between different FH subtypes and predict simvastatin efficacy and could effectively predict simvastatin doses, dosing intervals and duration required to achieve optimal simvastatin therapy.

During the last 20 years, various modeling approaches and software have been used to study different aspects of simvastatin pharmacokinetics (Moon and Smith, 2002; Ogungbenro et al., 2019; Tsamandouras et al., 2014, 2015; Kim et al., 2011; Methaneethorn et al., 2014; Lohitnavy et al., 2015; Kim et al., 2011; Wojtyniak et al., 2021), cholesterol metabolism (Paalvast et al., 2015; Wrona et al., 2015) and the effect of simvastatin on cholesterol levels (Kim et al., 2011). However, most of the work is difficult to validate or build upon due to the lack of accessibility of the models and software. Here, we provide an openly accessible, reproducible, and platform-independent whole-body model of simvastatin and LDL-C that facilitates reusability, extensibility, and comparability. The model was developed and validated on a large database of heterogeneous studies and is freely available in the open standard SBML (Keating et al., 2020).

The PBPK/PD model was able to accurately predict simvastatin pharmacokinetics and LDL-C reduction with simvastatin therapy. However, the model has several limitations. Most importantly, the model focused on the role of hepatic cholesterol synthesis and the LDLR pathway in plasma LDL-C. Because of the focus on the liver, the whole-body effect on cholesterol homeostasis was modeled only phenomenologically, e.g., dietary cholesterol uptake via an average uptake rate, and the role of other organs and tissues in systemic cholesterol homeostasis was not modeled in detail but lumped into an overall cholesterol consumption. Inhibition of HMG-CoA reductase by simvastatin metabolites was considered only in the liver, the major site of cholesterol synthesis, but the enzyme is also inhibited in other tissues, leading to potential side effects. While the LDLR pathway was modeled in some detail, cholesterol synthesis was simplified to the key steps of acetyl-CoA to HMG-CoA via HMG-CoA synthase and from HMG-CoA to cholesterol via HMG-CoA reductase.

An additional limitation of the model evaluation was the lack of data on hepatic cholesterol metabolism. Additional data other than plasma LDL-C concentrations would be very helpful to validate and improve the model.

The focus of the model was on LDL-C, but other substances such as VLDL-C, HDL-C and triglycerides also play an important role in hypercholesterolemia. Our data set already includes all data for these substances in simvastatin therapy, which were often reported together with LDL-C levels. Future work will extend the pharmacodynamic model to provide a broader view of changes in simvastatin therapy.

Importantly, we complement our open and accessible model with a large, open and accessible database of simvastatin pharmacokinetics and LDL-C pharmacodynamics in simvastatin therapy. The established database of simvastatin pharmacokinetics consists of pharmacokinetic studies with single or multiple doses of simvastatin in healthy patients. To our knowledge, no study has reported simvastatin pharmacokinetics in hypercholesterolemic patients. Therefore, it is unclear whether there is a systematic difference between simvastatin pharmacokinetics in healthy subjects and hypercholesterolemic patients.

Most of the reported data on the pharmacodynamics of simvastatin therapy, such as the reduction of plasma LDL-C, were poorly documented. This is consistent with our recent findings of poor quality pharmacokinetic data in the literature (Grzegorzewski et al., 2021, 2022). All pharmacodynamic studies reported baseline cholesterol levels, but data during and after simvastatin therapy were very heterogeneous. Some studies reported absolute changes in LDL-C, some relative changes, and some absolute plasma concentrations either with or without uncertainties such as standard deviation. The heterogeneity in reporting and the lack of complete data (concentrations, absolute changes, and relative changes) posed a major challenge for data integration. Additional information on the subjects was rarely and inconsistently reported (e.g. anthropometric information, diet). Most studies did not report the underlying cause of hypercholesterolemia.

Only a single study reported simvastatin pharmacokinetics and LDL-C pharmacodynamics(Loria et al., 1994). However, this study was limited to a single day and normocholesterolemic subjects. For model validation long-term studies measuring simvastatin pharmacokinetics and its LDL-C lowering effects in hypercholesterolemic patients would be a very valuable asset.

In this work, LDL-C lowering therapy with simvastatin in hypercholesterolemia was studied using a computational modeling approach. The main findings are: (i) hepatic LDLR turnover is highly heterogeneous among FH classes; (ii) despite this heterogeneity, simvastatin therapy results in a consistent lowering of plasma LDL-C independent of class; and (iii) simvastatin therapy shows a dose-dependent reduction in LDL-C. Our model suggests that the underlying cause of hypercholesterolemia in FH classes does not affect simvastatin therapy. Furthermore, our model supports the treatment strategy of stepwise dose adjustment to achieve target LDL-C levels. Both the model and the database are freely available for reuse.

## Data Availability

All data produced are available online at https://pk-db.com

## ACKNOWLEDGMENTS

FB, JG and MK are supported by the Federal Ministry of Education and Research (BMBF, Germany) within the research network Systems Medicine of the Liver (LiSyM, grant number 031L0054). FB and MK are supported by the German Research Foundation (DFG) within the Research Unit Programme FOR 5151 “QuaLiPerF (Quantifying Liver Perfusion-Function Relationship in Complex Resection - A Systems Medicine Approach)” by grant number 436883643. MK and HMT are supported by grant number 465194077 (Priority Programme SPP 2311, Subproject SimLivA). This work was supported by the BMBF-funded de.NBI Cloud within the German Network for Bioinformatics Infrastructure (de.NBI) (031A537B, 031A533A, 031A538A, 031A533B, 031A535A, 031A537C, 031A534A, 031A532B).

## AUTHOR CONTRIBUTIONS

FB and MK designed the study, developed the computational model, performed the analysis, and wrote the first draft of the manuscript. JG provided assistance with PK-DB (https://pk-db.com), data curation, and meta-analysis. All authors contributed to and critically revised the manuscript.

## DATA AVAILABILITY STATEMENT

The data sets analyzed in this study are available from PK-DB at https://pk-db.com.

---

**Table 1.**
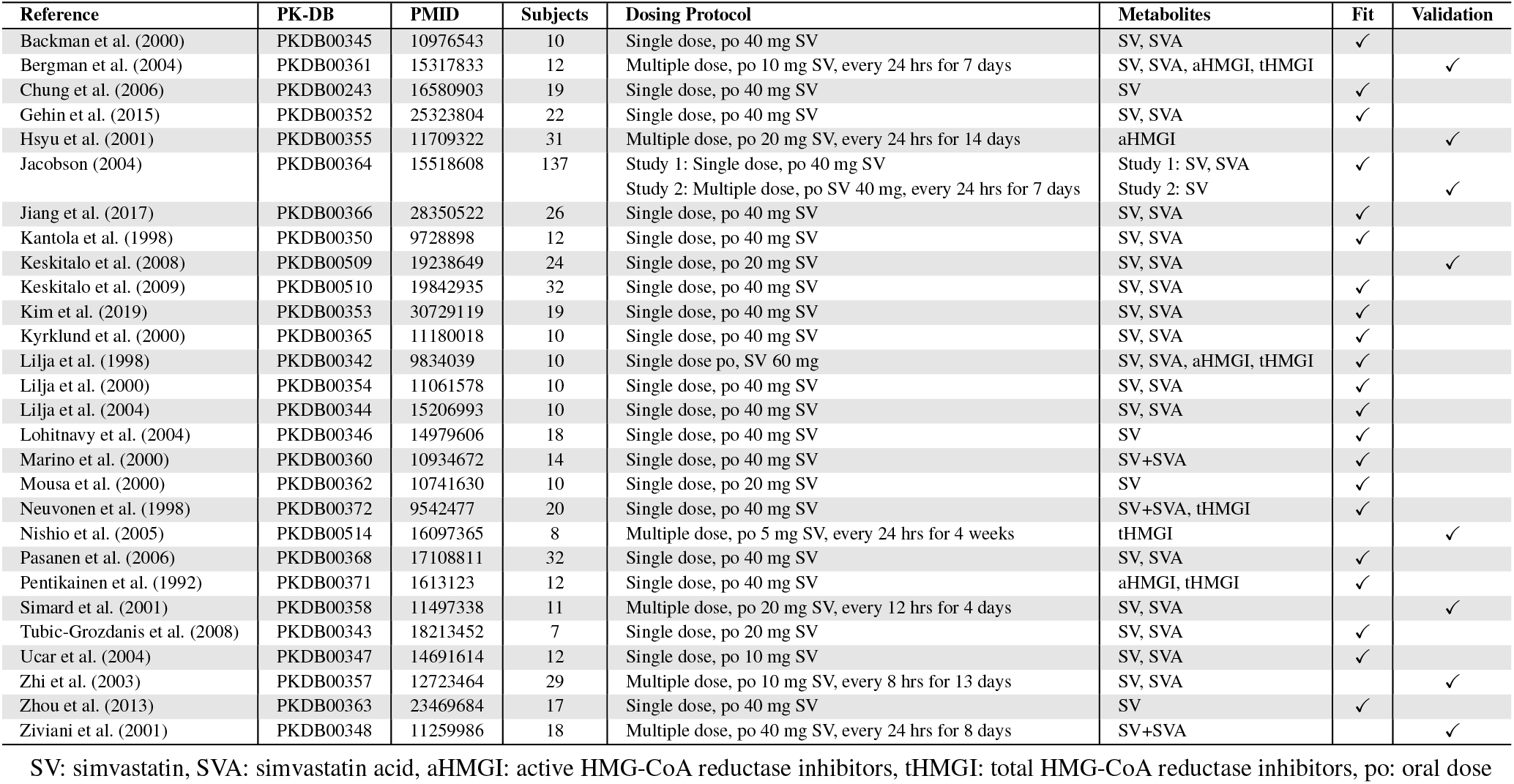
Overview of clinical studies with simvastatin pharmacokinetics.

**Table 2.**
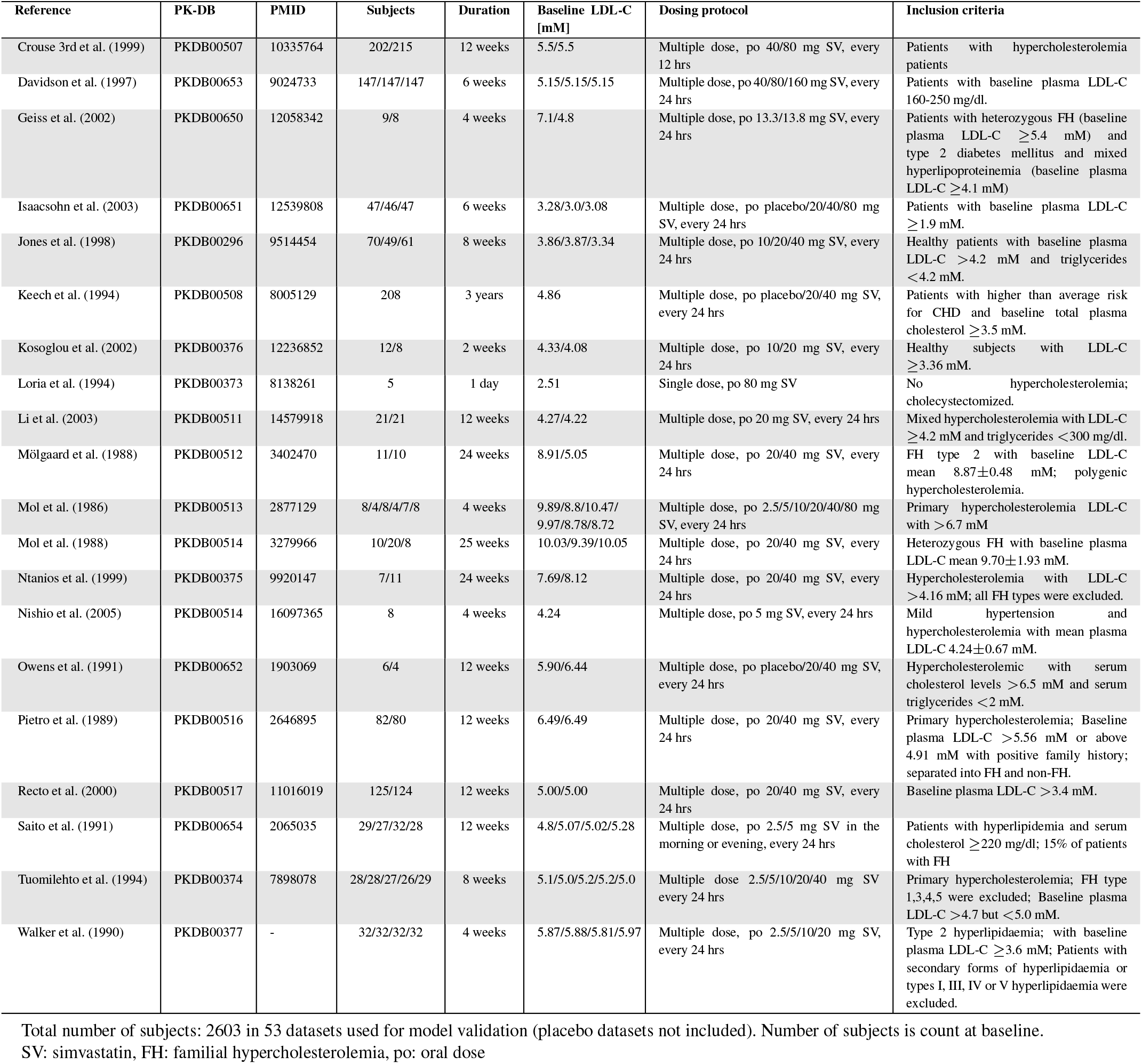
Overview of clinical studies with LDL-C measurements in simvastatin therapy.

