## Supplementary Material for "Simvastatin therapy in different subtypes of hypercholesterolemia – a physiologically based modelling approach"

### 1 SUPPLEMENTARY FIGURES

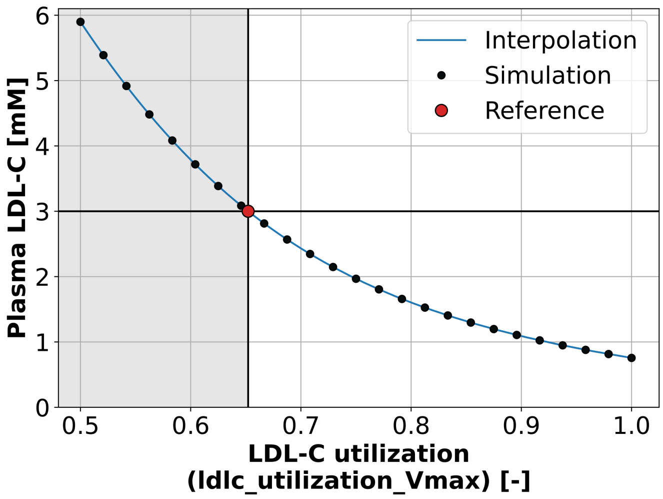


**Figure S1. Calibration curve for baseline LDL-C value.** The baseline plasma LDL-C concentration of the model was set to 3 mM using the LDL-C utilization parameter `ldlc_utilization_Vmax`. The model was simulated for 52 weeks and the steady-state plasma LDL-C concentration was determined for a given parameter value (black circles). The simulations were interpolated (blue curve) and the reference parameter corresponding to 3 mM plasma LDL-C was determined from the interpolation curve (red circle).

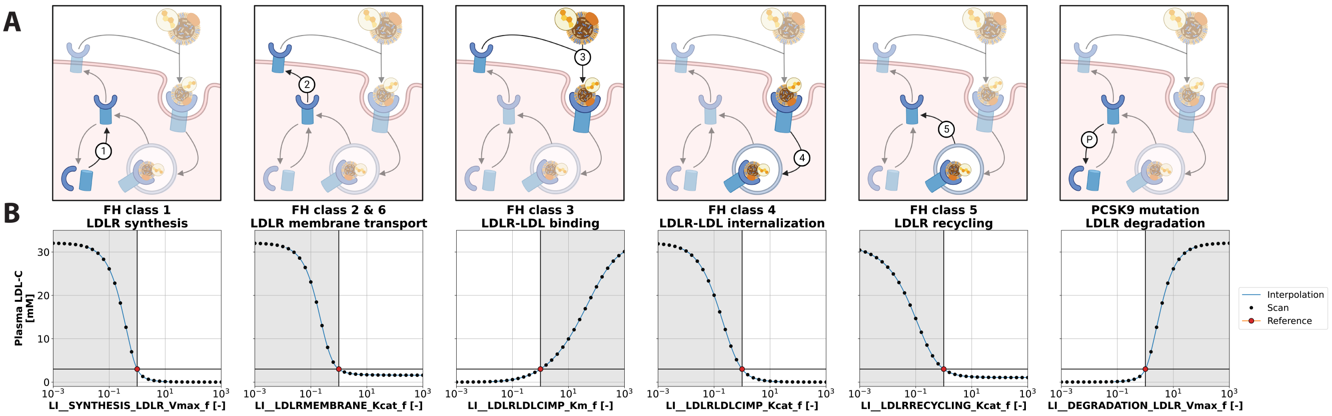


**Figure S2. Calibration curves for baseline LDL-C in hypercholesterolemia subtypes. A)** Graphic overview of the hypercholesterolemia subtypes: (1) LDLR synthesis; (2) transport of LDLR to the membrane; (3) binding of LDL-C to LDLR; (4) internalization of the LDLR-LDL-C complex; (5) recycling of LDLR; (P) degradation of LDLR; **B)** The model was simulated for 52 weeks and the steady-state plasma LDL-C concentration was determined for the given FH value (black circles). The simulations were interpolated (blue curve) and the interpolations were used to determine the parameter value corresponding to a given LDL-C level in hypercholesterolemia. Parameter values of 1.0 correspond to the reference model of 3 mM plasma LDL-C (red circle).

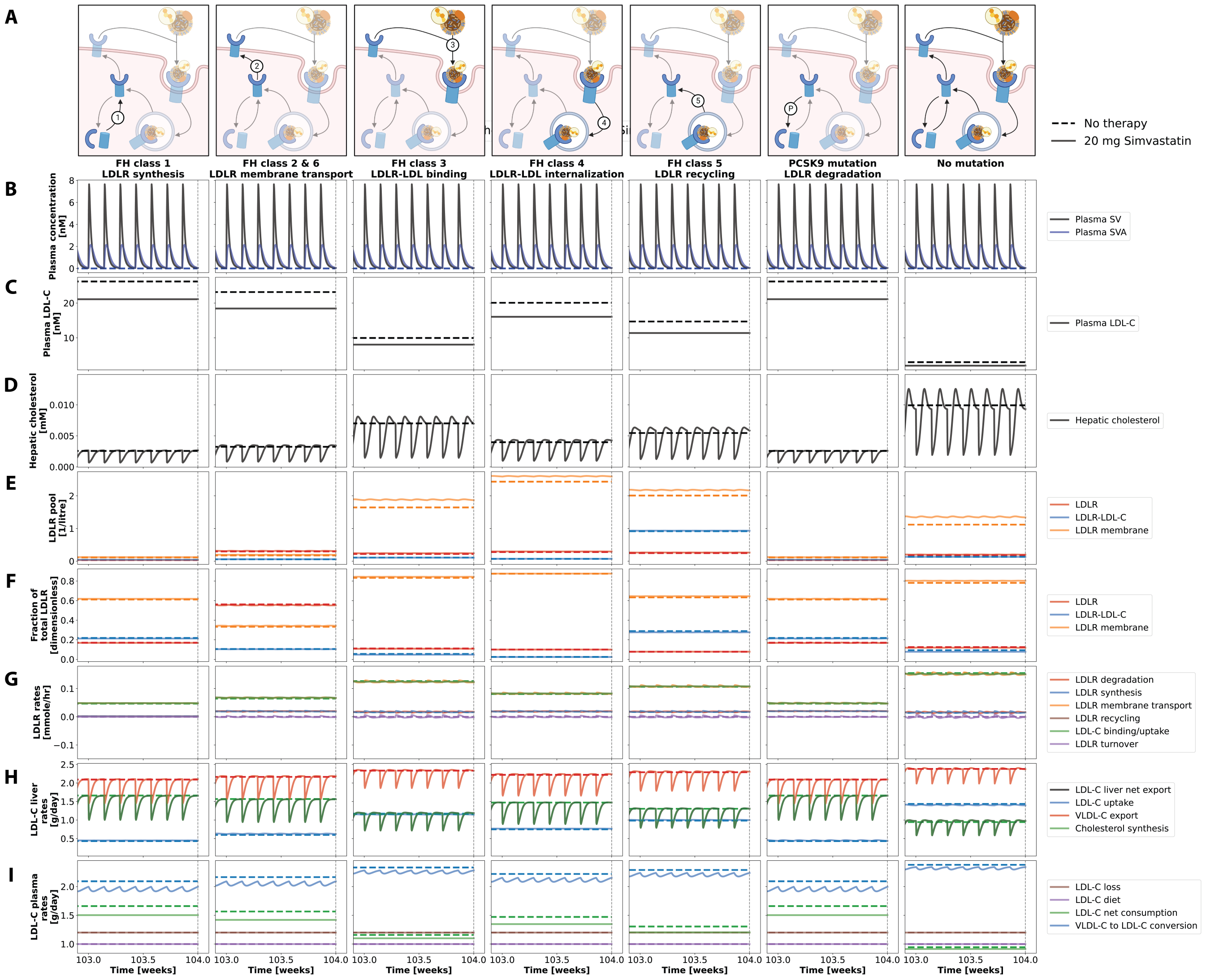


**Figure S3. Time course simulation of hypercholesterolemia classes and simvastatin therapy (last week).** The columns correspond to the different classes of hypercholesterolemia. Simulations as in Fig. 4, but only the last week is shown. **A)** Graphic overview of the hypercholesterolemia subtypes: no mutation in the LDLR pathway; (1) LDLR synthesis; (2) transport of LDLR to the membrane; (3) binding of LDL-C to LDLR; (4) internalization of the LDLR-LDL-C complex; (5) recycling of LDLR; (P) degradation of LDLR; **B)** Plasma concentration of SV and SVA. **C)** Plasma LDL-C. **D)** Hepatic cholesterol. **E)** Overview of the LDLR pool consisting of plasma LDLR, LDLR-LDL-C complex or membrane LDLR. **F)** Fractional LDLR pool. **G)** Rate of processes involved in LDLR turnover: LDLR degradation, LDLR synthesis, LDLR membrane transport, LDLR recycling, LDL-C binding/uptake, LDL-R turnover. **H)** LDL-C rates in the liver: LDL-C net export from liver, LDL-C uptake, LDL-C absorption, LDL-C export, cholesterol synthesis. **I)** Plasma LDL-C Rates: LDL-C fecal loss, LDL-C from diet, LDL-C net consumption, VLDL-C to LDL-C conversion.

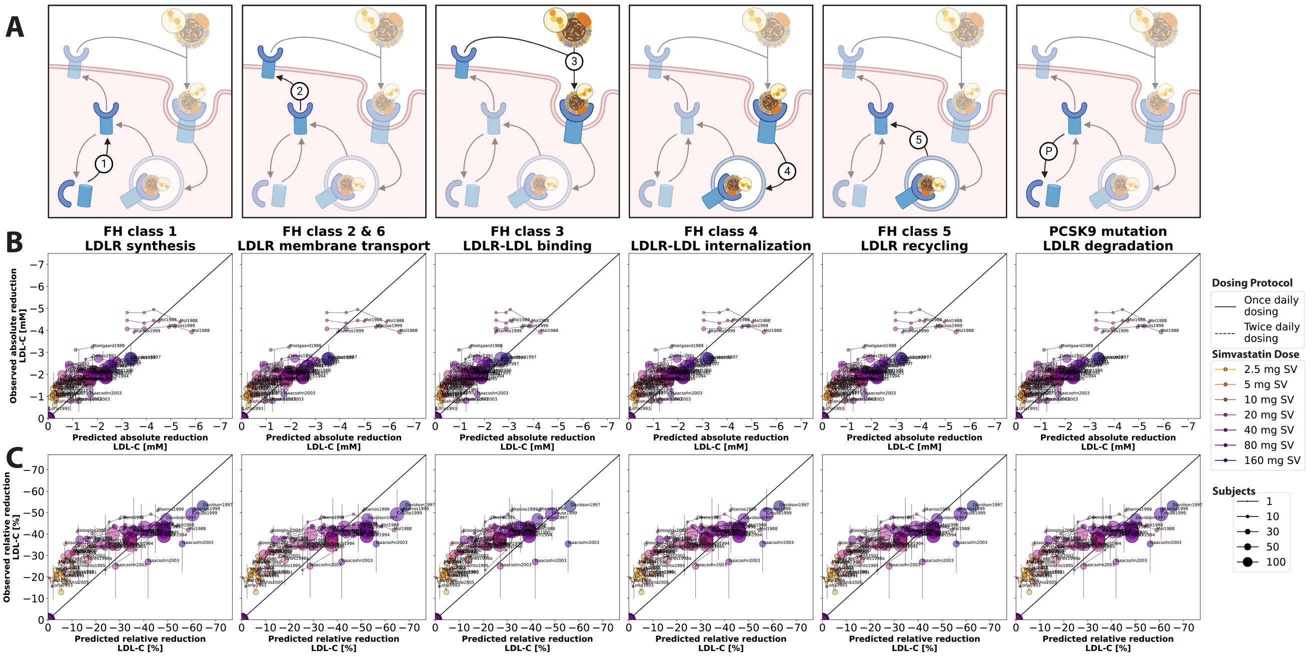


**Figure S4. Prediction of LDL-C change with simvastatin therapy by hypercholesterolemia subtype.** **A)** Graphic overview of the hypercholesterolemia subtypes: (1) LDLR synthesis; (2) transport of LDLR to the membrane; (3) binding of LDL-C to LDLR; (4) internalization of the LDLR-LDL-C complex; (5) recycling of LDLR; (P) degradation of LDLR; **B)** Observed vs. predicted absolute change in LDL-C. Observed changes are mean ± SD, predicted changes are for each hypercholesterolemia class. **C)** Same as (B) but for relative changes in LDL-C. Data from (Crouse 3rd et al., 1999; Davidson et al., 1997; Geiss et al., 2002; Isaacsohn et al., 2003; Jones et al., 1998; Keech et al., 1994; Kosoglou et al., 2002; Li et al., 2003; Loria et al., 1994; Mölgaard et al., 1988; Mol et al., 1986, 1988; Nishio et al., 2005; Ntanios et al., 1999; Owens et al., 1991; Pietro et al., 1989; Recto et al., 2000; Saito et al., 1991; Tuomilehto et al., 1994; Walker et al., 1990).

### 2 SUPPLEMENTARY TABLES

**Table S1.** Overview of physiological model parameters for the PBPK/PD model.

| Physiological parameter | Description | Value | Unit | Reference |
|---|---|---|---|---|
| BW | body weight | 75 | kg | ICRP (2002) |
| HEIGHT | body height | 170 | cm | ICRP (2002) |
| COBW | cardiac output per body weight | 1.25 | ml/s/kg | |
| HCT | hematocrit | 0.51 | dimensionless | Vander (2001); Herman (2016) |
| FVgi | fractional tissue volume gastrointestinal tract | 0.0297 | l/kg | Calculated based on (Jones and Rowland-Yeo, 2013; ICRP, 2002) |
| FVli | liver fractional tissue volume | 0.021 | l/kg | Jones and Rowland-Yeo (2013); ICRP (2002) |
| FVlu | lung fractional tissue volume | 0.0076 | l/kg | Jones and Rowland-Yeo (2013); ICRP (2002) |
| FVve | venous fractional tissue volume | 0.0514 | l/kg | Jones and Rowland-Yeo (2013); ICRP (2002) |
| FVar | arterial fractional tissue volume | 0.0257 | l/kg | Jones and Rowland-Yeo (2013); ICRP (2002) |
| FVpo | portal fractional tissue volume | 0.001 | l/kg | Jones and Rowland-Yeo (2013); ICRP (2002) |
| FVhv | hepatic venous fractional tissue volume | 0.001 | l/kg | Jones and Rowland-Yeo (2013); ICRP (2002) |
| FQgu | gut fractional tissue blood flow | 0.18 | dimensionless | Jones and Rowland-Yeo (2013) |
| FQki | kidney fractional tissue blood flow | 0.19 | dimensionless | Jones and Rowland-Yeo (2013) |
| FQh | hepatic fractional tissue blood flow | 0.215 | dimensionless | Jones and Rowland-Yeo (2013) |
| Mr_simva | molecular weight SV | 418.6 | g/mole | CHEBI:9150 |
| Mr_simacid | molecular weight SVA | 436.6 | g/mole | CHEBI:169041 |
| Mr_ldlc | molecular weight LDL-C | 386.654 | g/mole | PubChem CID:599 |
| LI__SYNTHESIS_HMG_Vmax | Synthesis Vmax for HMGR and HMGS | 0.001 | mmole/min/l | |
| LI__DEGRADATION_HMG_Vmax | Degradation Vmax for HMGR and HMGS | 0.0003 | mmole/min/l | |
| LI__DEGRADATION_LDLR_Vmax | Degradation Vmax for LDLR | 0.0006 | mmole/min/l | |
| LI__VLDLCEXP_Vmax | VLDL-C export from liver Vmax | 2.5 | dimensionless | Scaled to 2.5 mg/day |
| LI__VLDLCEXP_Km_cho | VLDL-C export from liver Km depending on hepatic cholesterol | 0.0005 | mmol/l | Assumed to be have high dependency on hepatic cholesterol |
| LI__LDLRLDLCIMP_Kcat | LDLR-LDLC binding and internalization Kcat | 6 | dimensionless | Scaled to 1.0 mg/day |
| LI__LDLRLDLCIMP_Km | LDLR-LDLC binding and internalization Km | 3 | mmol/l | Based on assumption that plasma LDL-C is 3 mM |
| LI__LDLRMEMBRANE_Kcat | Transport of hepatic LDLR to the membrane Kcat | 0.025 | mmole/min/l | (Goldstein and Brown, 1990) |
| LI__LDLRRECYCLING_Kcat | Recycling of hepatic LDLR Kcat | 0.0125 | mmole/min/l | (Goldstein and Brown, 1990) |
| LI__HMGCOASYNTHASE_Kcat | HMGS Kcat | 245 | l/mmol | Assumption higher than LI__HMGCOARECUCTASE_Kcat |
| LI__HMGCOAREDUCTASE_Kcat | HMGR Kcat | 0.7 | dimensionless | Scaled to 0.7 mg/day (Brody, 1999) |
| LI__HMGCOAREDUCTASE_Km | HMGR Km | 0.004 | mmol/l | (Istvan and Deisenhofer, 2001) |
| LI__HMGCOASYNTHASE_Ki_hmgcoa | Inhibition Constant HMG-CoA on HMGS | 0.022 | mmol/l | Scaled on hepatic HMG-CoA concentration (Corkey et al., 1988) |
| LI__HMGCOAREDUCTASE_Ki_simacid | Inhibition constant of SVA on HMGR | 1.5E-6 | mmol/l | (Corsini et al., 1995; Istvan and Deisenhofer, 2001) |
| LI__simmets_inhibitor_activity | Inhibitory activity of SVM in relation to SVA | 0.5 | dimensionless | |
| LI__HMG_kr_cho | Protein activity regulation Kr for HMGR and HMGS by hepatic cholesterol | 0.02 | mmol/l | Assumptiom based on hepatic cholesterol concentration (Chuang et al., 2017) |
| LI__LDLR_kr_cho | Protein activity regulation Kr for LDLR by hepatic cholesterol | 0.02 | mmol/l | Assumption based on hepatic cholesterol concentration (Chuang et al., 2017) |
| ldlc_utilization_Vmax | Vmax plasma LDL-C utilization | 0.6521 | dimensionless | see Fig. S1 |
| ldlc_diet_k | Daily dietary cholesterol uptake k | 1 | dimensionless | Scaled to 1 mg/day (Brody, 1999) |
| ldlc_loss_k | Daily cholesterol loss into feces k | 1.2 | dimensionless | Scaled to 1.2 mg/day (Cohen, 2008) |
| ldlc_consumption_Km | Consumption and usage of cholesterol in the body Km | 60 | mmol/l | Assumption small dependency on plasma LDL-C |
| vldlc_conversion | Conversion rate of plasma LDLC into VLDLC | 2 | l/mmole | Scaled to 2 mg/day |

LDL-Cholesterol: LDL-C; VLDL-Cholesterol: VLDLC; LDL-receptor: LDLR

**Table S2.** Overview of optimized parameter using parameter fitting for the simvastatin model. Parameters are rounded to four digits.

| Fit parameter | Description | Value | Unit | Fitted |
|---|---|---|---|---|
| ftissue_simva | SV tissue distribution rate | 0.1502 | l/min | ✓ |
| ftissue_simacid | SVA tissue distribution rate | 38.5622 | l/min | ✓ |
| Ka_dis_simva | SV dissolution rate | 0.3582 | 1/hr | ✓ |
| SIMVA_CYP3A4LI_Vmax | $V_{max}$ of the reaction of SV and SVA to SVM in the liver | 0.1084 | mmole/min/l | ✓ |
| GU__SIMVAABS_k | rate of absorption of SV in the intestine/gut | 1.5088 | 1/hr | ✓ |
| GU__SIMMETSABS_k | rate of absorption of SVM in the intestine/gut | 0.0225 | 1/hr | ✓ |
| GU__F_abs_simmets | fraction absorbed SVM in the intestine | 0.2691 | dimensionless | ✓ |
| GU__SIMVA_CYP3A4_f | scaling of intestinal Vmax CYP3A4 relative to liver for SV | 0.0010 | dimensionless | ✓ |
| GU__SIMVAEXP_f | factor for SV export from the intestine/gut | 10.9627 | dimensionless | ✓ |
| GU__SIMMETSEXP_f | factor for SVM export from the intestine/gut | 29.2759 | dimensionless | ✓ |
| LI__SIMACID__CYP3A4_f | factor CYP3A4 catalyzed reaction of SVA to SVM in the liver | 4.7122 | dimensionless | ✓ |
| LI__SIMVAIMP_f | factor of SV import in the liver | 3.2829 | dimensionless | ✓ |
| LI__SIMACIDEXP_f | factor of SVA export from the liver | 0.0081 | dimensionless | ✓ |
| LI__SIMMETSEXP_f | factor of SVM export from the liver | 21.572 | dimensionless | ✓ |
| LI__ESTERASE_f | factor of esterase catalyzed reaction of SV to SVA in the liver | 0.1696 | dimensionless | ✓ |
| LI__EHC_simmets_k | rate of SVM transport into the bile | 3.1138 | 1/min | ✓ |
| KI__SIMMETSCL_k | rate of SVM clearance from the kidney into the urine | 1.5712 | 1/min | ✓ |

SV: simvastatin, SVA: simvastatin acid, SVM: simvastatin metabolites, CYP3A4: cytochrome P450 3A4

**Table S3.** Overview of FH scaling parameters to simulate FH in different classes. These are by default 1, which represents normal functioning LDLR pathway reactions.

| FH parameter | Description | Scan | Unit |
|---|---|---|---|
| LI__SYNTHESIS_LDLR_Vmax_f | LDLR synthesis (FH Class 1) | [0.01, 100] | dimensionless |
| LI__LDLRMEMBRANE_Kcat_f | LDLR membrane transport (FH Class 2 & 6) | [0.01, 100] | dimensionless |
| LI__LDLRLDLCIMP_Km_f | LDLR-LDL binding (FH Class 3) | [0.01, 100] | dimensionless |
| LI__LDLRLDLCIMP_Kcat_f | LDLR-LDL internalization (FH Class 4) | [0.01, 100] | dimensionless |
| LI__LDLRRECYCLING_Kcat_f | LDLR recycling (FH Class 5) | [0.01, 100] | dimensionless |
| LI__DEGRADATION_LDLR_Vmax_f | LDLR degradation (PCSK9 mutation) | [0.01, 100] | dimensionless |

LDL-Cholesterol: LDL-C; VLDL-Cholesterol: VLDLC; LDL-receptor: LDLR
